# AURORA: Analysing and understanding responses to oncological regimens with artificial intelligence

**DOI:** 10.64898/2026.08.30.26361778

**Authors:** Adrian Lebmeier, Tim Lindner, Christian Karl, Thorsten Schöler, Andreas Rank

## Abstract

**Background:** Immunochemotherapy (ICT) is considered standard in regards to care for small-cell lung cancer (SCLC) in extensive stages, yet reliable biomarkers for treatment response remain elusive. While previous univariate analyses suggest specific peripheral lymphocyte subsets correlate with survival, the systemic immune response involves complex, multivariate interactions that require advanced analytical approaches.

**Methods:** This paper analysed high-dimensional flow cytometry data from 32 patients with stage IV SCLC treated with carboplatin, etoposide, and atezolizumab. Peripheral blood was analysed at baseline (**V**_0_) and longitudinally during treatment. To identify potential early predictive biomarkers and mitigate sample attrition in later cycles, we focused on baseline and measurements after two cycles of ICT (**V**_1_). We employed a rigorous machine learning framework utilising nested cross-validation, bootstrapping, and permutation-based statistical testing to evaluate eleven different regression and survival models.

**Results:** Under model-appropriate metrics, regressors did not generalise (*R*^2^ *<* 0); conversely, censoring-aware Random Survival Forests (RSF) successfully extracted robust prognostic signatures. Baseline immune profiles (**V**_0_) achieved a concordance index (C-index) of 0.66 (*p* = 0.015), while dynamic changes from **V**_0_ to **V**_1_ (**ΔV**) achieved a C-index of 0.65 (*p* = 0.022). Crucially, absolute values measured after two cycles of ICT (**V**_1_) yielded no significant signal (*p* = 0.445). Feature importance analysis confirmed the prognostic value of Th17 normalisation and identified Naive Regulatory T cells and Memory B cells as candidate components.

**Conclusion:** Machine learning validation confirms a predictive signal in the peripheral immune profile of SCLC patients. Early dynamic shifts in the balance between regulatory and effector immune arms are associated with prognosis, contrasting with the lack of signal in absolute counts after two cycles of ICT. These findings establish a proof of concept for multivariate liquid biopsy immune profiling, warranting confirmation in larger cohorts and highlighting the necessity of integrating systemic and tumour-intrinsic data.

## Introduction

SCLC is an aggressive malignancy, accounting for approximately 15 % of all lung cancers [1] and typically diagnosed at advanced stages. It is characterised by a short doubling time, rapid growth, and the early occurrence of widespread metastases. Historically, median overall survival for extensive-stage disease was only 9 to 10 months [2]. Currently, there are no curative treatment options for patients with extensive-stage (UICC stage IV [3]) SCLC. The introduction of Immunochemotherapy (ICT), a combination of first-line platinum-based chemotherapy and immune checkpoint inhibition (CPI) with anti-PD-L1 (anti-programmed death ligand 1) antibodies such as atezolizumab or durvalumab, has modestly improved median overall survival by roughly two months to around 12 to 13 months [4, 5]. Despite these advances, reliable predictive biomarkers for ICT response in SCLC remain unclear. Classical tumour-intrinsic markers, such as PD-L1 expression [6] and tumour mutational burden (TMB) [7], which are predictive in non-small-cell lung cancer (NSCLC), have consistently failed to correlate with treatment response in SCLC [4, 5]. This highlights the urgent need for immune-based biomarkers that reflect the patient’s **systemic immunological status rather than solely tumour-intrinsic features**.

The success of CPI relies heavily on reactivating the patient’s immune response, particularly cytotoxic T lymphocytes. Therefore, the peripheral blood, as an easily accessible compartment, offers a potential window into the systemic anti-tumour immune status and the effectiveness of CPI. The systemic immune environment in SCLC is characterised by profound immune suppression and dysregulation of peripheral lymphocyte subsets. Schmälter et al. [8] established a baseline immune profile in stage IV SCLC patients, observing significantly decreased peripheral B and Natural Killer (NK) cell counts in comparison to healthy controls. This is consistent with findings in other solid tumour entities. Longitudinal monitoring during ICT showed therapy-associated shifts: significantly increasing exhausted CD4+ and CD8+ T cells, continuing decline in B cells, and dynamically changing Th subsets. Critically, **a decrease in Th17 cells after two cycles of ICT was significantly associated with prolonged overall survival**(*p* = 0.006), suggesting that the normalisation of this specific subset could serve as a promising early predictive biomarker for treatment efficacy. Building on these findings, the present study is designed to systematically assess the entire peripheral lymphocyte panel to identify multivariate prognostic signatures and addresses two primary hypotheses regarding patient overall survival (OS):

*H*_1_ **Baseline Predictors**: The baseline composition of peripheral blood lymphocyte subsets prior to the initiation of ICT is associated with OS.
*H*_2_ **Dynamic Early Immune Signatures**: Dynamic changes in the composition of peripheral blood lymphocyte subsets, expressed as the absolute difference from baseline after two cycles of ICT, are associated with OS.

To address these hypotheses and to differentiate robust, high-performance prognostic signatures from spurious correlations, this study evaluates an established pool of machine learning models that are widely applied in clinical prediction tasks, particularly given the often high-dimensional, heterogeneous nature of clinical data [9–12], as well as a model with promising potential on small datasets [13]. We employed regression predictors (XGBoost, Random Forest, Ridge, Lasso, Support Vector Regression, k-Nearest Neighbour, and TabPFN) and specialised survival models tailored for right-censored data (Random Survival Forest, penalised Cox regression, classical CoxPH, and Fast Survival SVM). This methodology provides a systematic evaluation of predictive performance, stability, and robustness of the models, thereby identifying models that consistently achieve high predictive accuracy and reliable performance for clinical outcomes based on immunological data. This approach is further described in detail in Models and Approach. **Sample Strategy and Attrition:** To identify potential early predictive biomarkers and mitigate sample attrition in later cycles, this study focused on baseline and measurements after two cycles of ICT (**V**_1_). We recognise that complete-case pairing can introduce selection bias; therefore, we frame this analysis as proof of concept, acknowledging that missing-data-aware sensitivity analyses would be required for definitive confirmation.

## Materials and Methods

### Dataset

The dataset comprises 32 individuals in UICC stage IV small-cell lung cancer (SCLC). Flow cytometry is used to represent individual peripheral blood lymphocyte subsets during treatment with ICT. A topological overview is shown in Fig 1. The dataset restricted to the variables analysed in this study is included in S3 Table.

**Fig 1.**
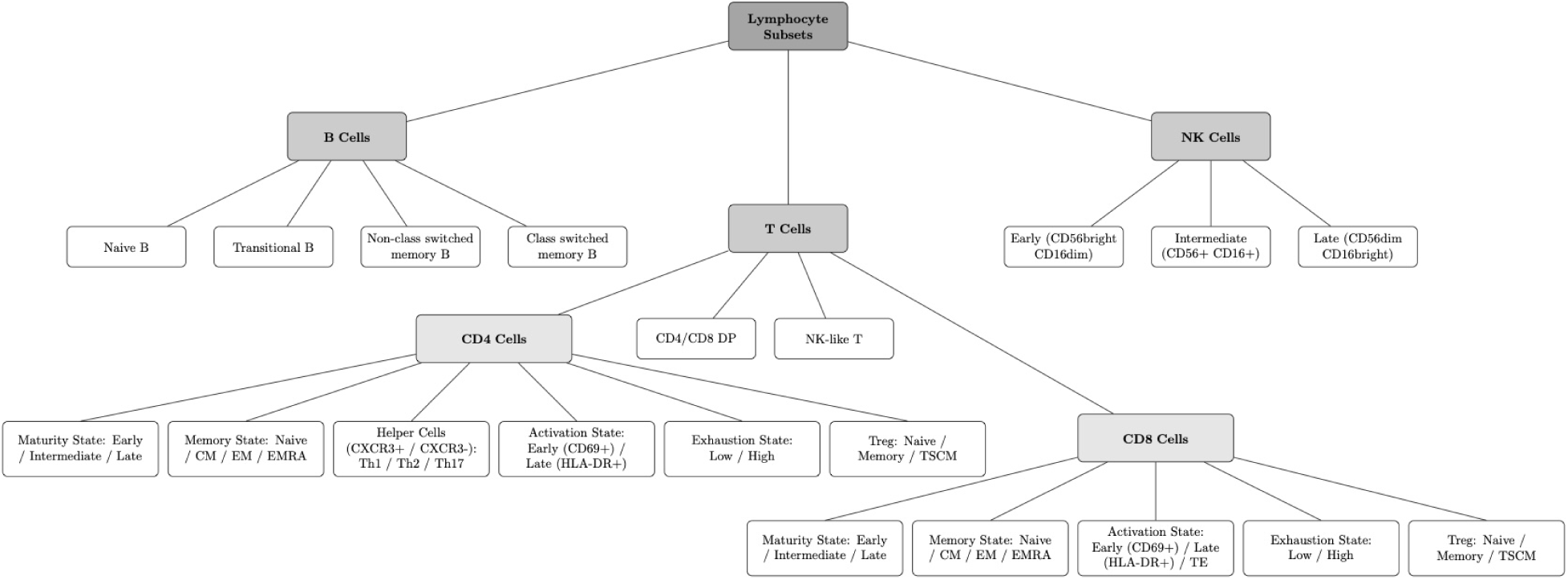
Lymphocyte subsets in peripheral blood analysed by immunophenotyping via flow cytometry. Main subpopulations include B cells, T cells (CD4 cells versus CD8 cells), and NK cells. CM centre memory, CXCR3 C-X-C motif chemokine receptor 3, DP double positive, EM effector memory, EMRA effector memory with RA expression, NK natural killer, TE terminal effector cells, Th T helper cells, and TSCM T stem cell-like memory.

The measurements were repeated several times and were noted as different therapy states. **V**_0_ indicates pre-ICT and **V**_1_ represents the patient’s immune cell subset values after two cycles of ICT, respectively. The original dataset, described in [8] includes twelve therapy cycles (**V**_0_ – **V**_6_ measured every two therapy cycles) as well as healthy individuals used for uni- and multivariate analysis. Since the already small dataset size decreases over time (*<* 10 samples after **V**_2_) and the research questions in this study specifically addresses **V**_0_ and **V**_1_ stages, corresponding columns after **V**_1_ and the healthy control group were excluded.

The tabular dataset was further divided into three individual subsets that were analysed in isolation. Columns that are foundational for the analysis were included in each subset and are further referred to as base columns. These include age and gender, as well as the separated target column overall survival (OS). Last-follow-up (LFU) was dropped for non-survival models. For survival analysis, OS and LFU were processed and included as the columns time and event, respectively, where time states the OS and event signals whether the event has occurred (1) or not (0). In the latter case, the data is censored which describes that the survival time is only known through the last follow-up meaning that a patient has an OS of at least the LFU time. Three out of 32 are censored. The median age is 70 years (44 – 83) and is constituted of 41 % female and 59 % male participants. The mean OS is 279.41 days with a standard deviation of 209.93 (13 – 757).

The first subset only includes patients before ICT (**V**_0_) and addresses the first research question *H*_1_. After removal of missing values, it comprises 24 patients (samples) and 56 immune cell subsets (columns or features). The second subset, named **V**_1_, includes data only from patients after two cycles of immunotherapy with data including 20 patients and 56 features. The third dataset comprises delta values of the aforementioned subsets and were calculated as follows:

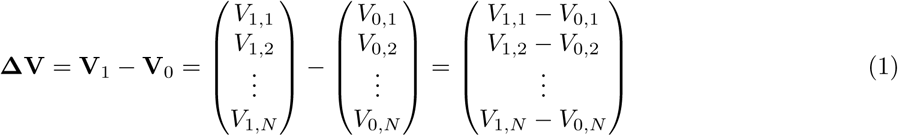

where:

**ΔV** is the resulting **Delta Feature Vector**, comprising the element-wise differences for all patients

**V**_0_ is the **Baseline Vector**, representing the set of measured values for all patients before therapy.

**V**_1_ is the **Post-Treatment Vector**, representing the set of measured values for all patients after two cycles of ICT.

*N* is the **total number of patients**, in the study.

*V_t,i_* is a **scalar value** representing the measured feature for an individual patient *i* at time point *t* (where *t* ∈ {0, 1} and i ∈ {1, . . . , *N*}).

The resulting subset **ΔV** describes dynamic changes in the subsets and have resulted in findings in the previous paper [8]. Due to mismatched missing values in either **V**_0_ or **V**_1_, the sample size was reduced to 16 patients with 56 features. The next section describes how the full dataset as well as the described subsets are used to conduct exploratory data analysis (EDA), and data pre-processing according to the models’ individual prerequisites.

### Ethics Statement and Data Access

The study was conducted according to the guidelines of the Declaration of Helsinki, and approved by the Institutional Review Board of the University Hospital Augsburg, Germany (protocol code 2020-10, approved 25th April 2020). Written informed consent was obtained from all patients and healthy control participants involved in this study. The data were accessed for research purposes between 22 April 2025 and 19 January 2026. During data collection and extraction, authors affiliated with the University Hospital Augsburg (Rank, A.) had access to information that could identify individual participants. The data were pseudonymised before being transferred to the computational authors, who had no access to identifying participant information.

## Methodology

This section describes the overall methodological framework used to address the research objectives introduced above. The framework was designed by guidance of the *Cross-Industry Standard Process for Data Mining (CRISP-DM)* [14] and aims to ensure a reproducible, unbiased, and interpretable analysis of the medical cancer dataset.

An overview of the methodology is shown in Fig 2. Initially, the dataset is inspected for structure and quality through exploratory data analysis (EDA), including skewness, correlations, and detection of missing values, outliers, and duplicate samples. Categorical data (gender) are one-hot encoded, and missing values are dropped, since imputation does not improve results (see Pre-processing). The data is then organised into three subsets as described in the Dataset section. The cleaned datasets are then fed into a variety of machine learning models and further split into folds or subsamples using nested repeated k-fold cross-validation (CV) and bootstrapping. To reduce high-dimensionality and overfitting, a defined set of pre-processing steps and filters is applied and tested in each fold and bootstrap, respectively. The inner CV is used for hyperparameter optimisation, as further described in Cross-Validation. After CV, the best pre-processing steps and model hyperparameters stored during folds are used to run bootstrapping with this pre-processing and parameter setup. This is done for each of the three subsets. A CV run is defined as one particular filter setup for a single data subset, including all repetitions and nested folds, each yielding an intermediate result. A bootstrap run is defined as a complete set of iterations using the filter setup and model hyperparameters that performed best in the corresponding CV run, applied to each data subset.

**Fig 2.**
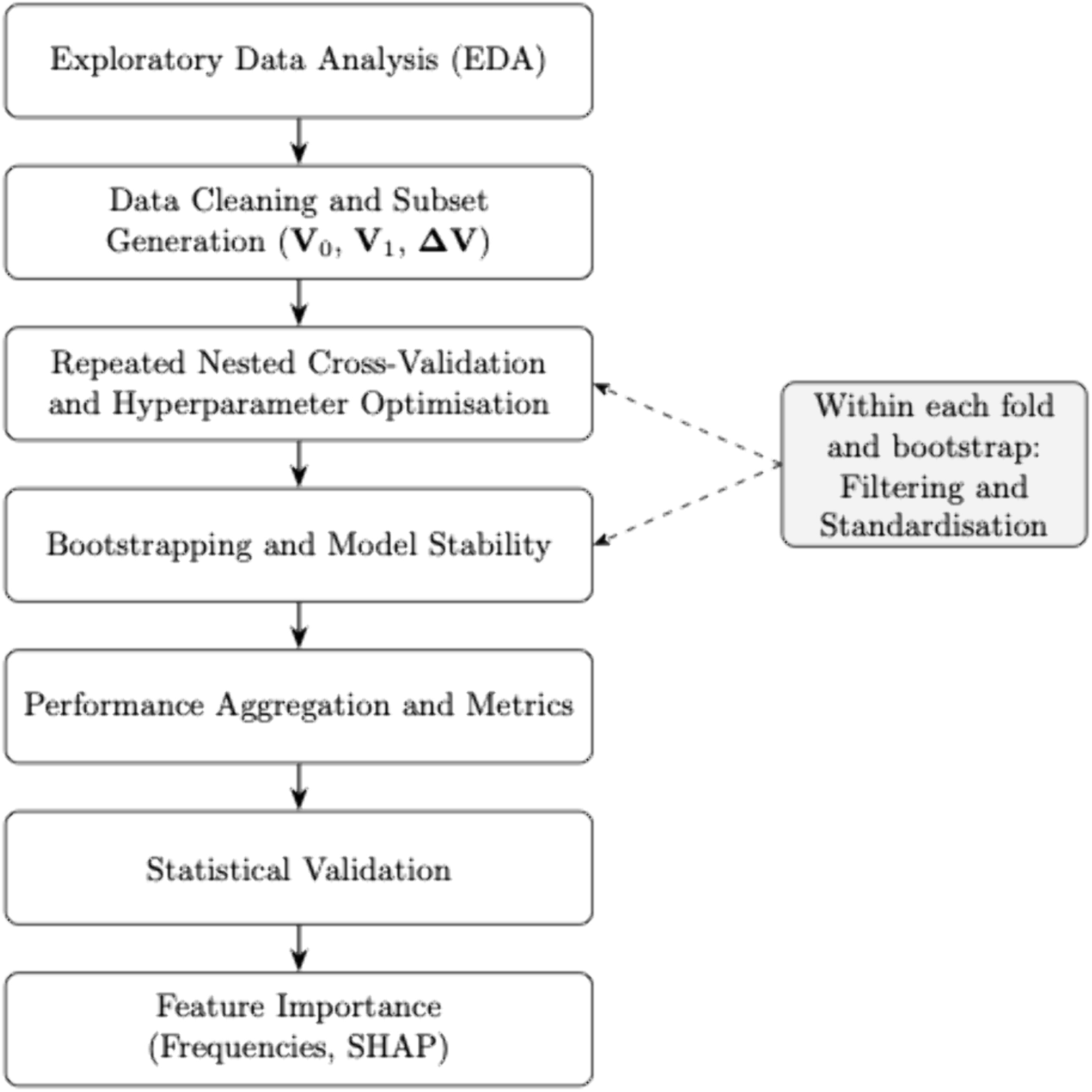
Overview of the methodological framework.

To assess whether the models captured meaningful signals rather than fitting noise, permutation-based statistical tests were implemented on the best models after all runs. Two approaches were applied. The first compared the model’s performance (C-index) against a null distribution, resulting in a calculated p-value. The second addressed the evaluation of whether the predictive performance of the best model per dataset differed significantly between the datasets **V**_0_ and **ΔV**. This is further described in Statistical Significance and Model Comparison.

After each CV run, a SHapley Additive exPlanations (SHAP) explainer is fitted on the **full dataset using the best model from this run**, solely to interpret which features may contribute to the target OS. SHAP provides feature-level attributions that fairly distribute a model’s prediction among features, capturing each feature’s average contribution across all possible subsets [15]. After each bootstrap run a stability-statistics table is generated, ranking the most stable features and their importance for model accuracy using either the model’s internal importance scores (e.g. XGBoost splitting) or permutation importance for other models. Additionally, after each bootstrap run, a SHAP explainer is trained on the full dataset for interpretability. During each run, performance metrics are computed for each fold or bootstrap iteration, and the mean values as well as 95 % confidence intervals are determined. This procedure is applied to each model—regular and survival, respectively. After all runs are completed, the best models per model type (regular or survival) and per data subset are summarised in a single table. The summary includes SHAP plots and feature importance tables of the best regular and survival models from the CV and bootstrap runs, respectively.

### Frameworks and Tools

We used Python as the main programming language and environment due to its extensive frameworks well suited for data science project. For automated and comparable training and evaluation, we used Kedro [16], an open-source data science framework. Other prominent frameworks are scikit-learn [17] and XGBoost [18], from which a variety of models were used, as well as the Python libraries pandas [19] and NumPy [20] for data processing. For survival models, we chose to use the frameworks scikit-survival [21] and the survival analysis library lifelines [22].

### Exploratory Data Analysis

Exploratory data analysis (EDA) was conducted to gain an initial understanding of the dataset and to inform subsequent preprocessing steps. Feature columns were renamed according to therapy cycles, control subjects were removed and data subsets were created as described in Dataset. Descriptive statistics were used to assess data quality and distribution, indicating non-normal distributions, the presence of missing values, and no duplicates. Collinearity was evaluated using Spearman correlation matrices for each subset. Outcome sparsity and skewness were analysed using a binned histogram of OS values (Fig 3), which showed a bi-modal distribution with an early peak, a long right tail (mean *>* median), and limited data in higher survival ranges (*>* 800). Feature-wise scatter and box plots further indicated the presence of outliers. Dimensionality reduction techniques, e.g. the factor analysis or principal component analysis (PCA), were explored during EDA but not pursued further, as preserving the interpretability and identification of individual relevant features was prioritised over maximal predictive performance. The insights gained from EDA guided the preprocessing strategy described in the following subsection.

**Fig 3.**
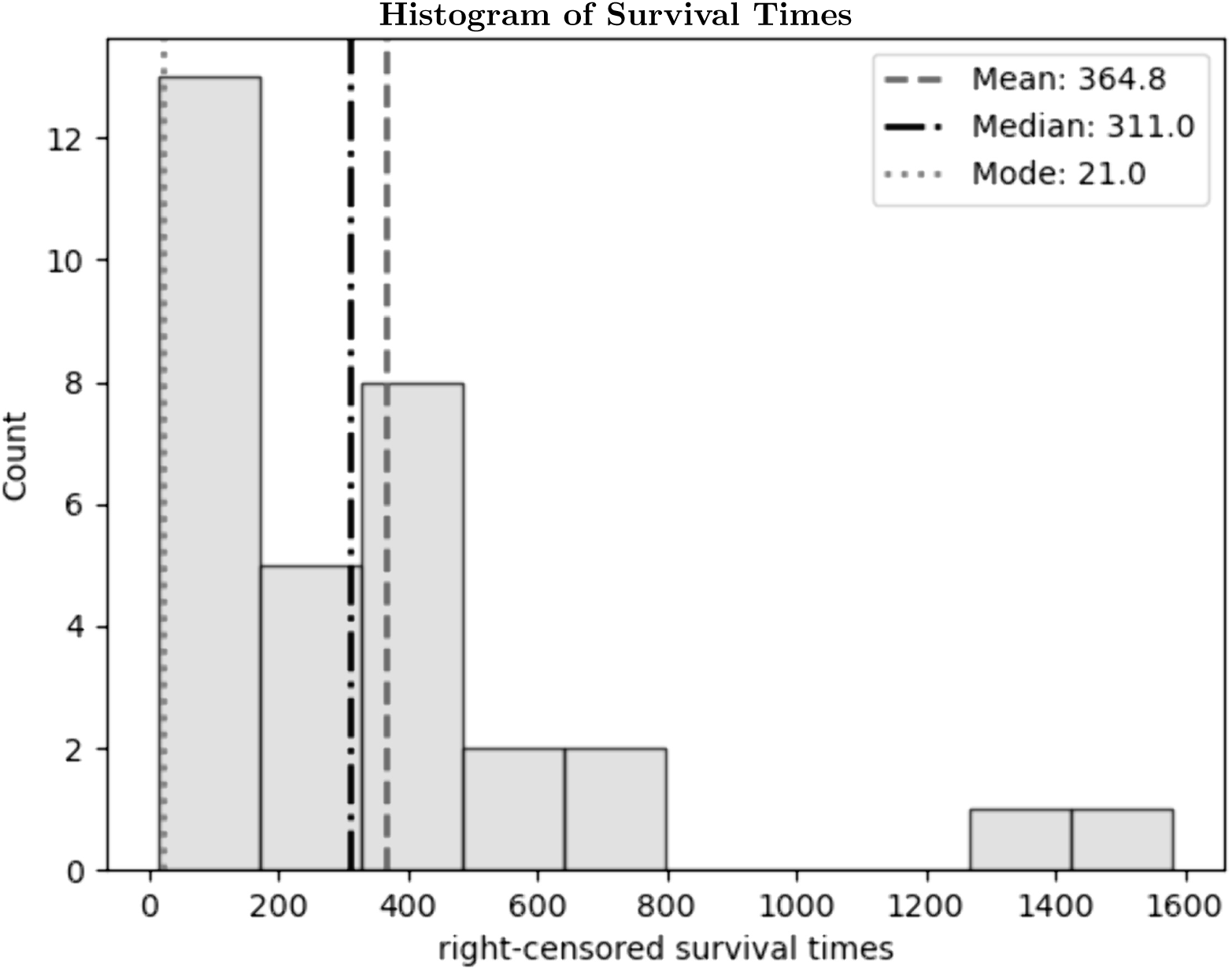
Histogram of survival times including overall survival (OS) and censored times (LFU) with mean, median, and mode.

### Histogram of Survival Times

### Pre-processing

The original dataset contained missing values across therapy cycles. KNN-based and domain-informed imputations were tested, but as these did not improve model performance, rows with missing values were removed for all subsequent analyses.

To reduce dimensionality and mitigate overfitting, feature filtering and standardisation were applied within each cross-validation and bootstrap fold. Highly correlated features were identified using Spearman correlation and removed, while features with weak univariate associations to the target (*p >* 0.05) were also filtered. Continuous features were standardised, resulting in a zero mean and unit variance, whereas categorical features (e.g. gender) were one-hot encoded.

The impact of pre-processing on model performance was explored by **testing all combinations of filtering and standardisation steps**. All combinations of pre-processing steps were explored to evaluate their impact on model performance. Full technical details and thresholds are provided in Supporting information.

### Experimental Setup

#### Cross-Validation

Cross-validation is a resampling procedure used to estimate model generalisation and select hyperparameters in a data-efficient and unbiased way [23].

In this work, we employed a nested cross-validation scheme. The inner loop for hyperparameter optimisation utilised GridSearchCV, while the outer loop for performance estimation employed RepeatedKFold. Both were implemented using the *scikit-learn* library [17].

This approach has been shown to provide more reliable and less biased performance estimates, particularly in small-sample, high-dimensional studies where conventional single CV runs tend to produce overly optimistic results [24].

Each run used a 3-fold outer loop for unbiased performance estimation and a 3-fold inner loop for hyperparameter tuning. The outer procedure was **repeated** 58 **times** (3 · 58 = 174 outer test folds in total). Within each outer split, the inner loop performed **repeated** 3**-fold CV with** 20 **repeats** (3 · 20 = 60 inner resamples) and conducted grid-search hyperparameter optimisation on the training portion only. To avoid excessive model flexibility given the limited sample size, all hyperparameter grids were deliberately kept narrow and restricted to commonly used values. The complete grids explored for each model are provided in the Supporting information.

For regular (non-survival) models, the model-selection metric was *R*^2^ [25], and for survival models, it was Harrell’s C-index (Concordance Index). The configuration that maximised the inner-loop metric was refit on the full inner-training data (outer train fold) and evaluated once on the held-out outer test fold. All data preprocessing steps were **fit exclusively on the corresponding training partition** within each inner/outer split and then applied to the validation/test partition to avoid data leakage.

Across the 174 outer-fold evaluations, we report the mean and 95 % confidence intervals computed using Student’s *t*-distribution. These were implemented in Python using the *SciPy* library [26]. For a set of outer test scores 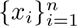, the two-sided (1 *− α*) confidence interval is:

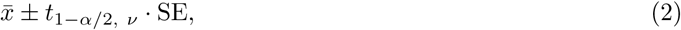

where:

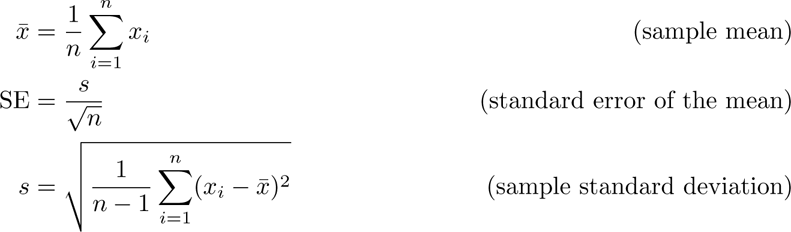

and *t*_1_*_−α/_*_2,_ *_ν_* is the critical value of Student’s *t*-distribution with *ν* = *n* − 1 degrees of freedom.

For a 95 % confidence level (*α* = 0.05), this approach accounts for finite-sample uncertainty and yields wider intervals than the normal-approximation approach (which assumes a known population standard deviation), making it preferable when the sample size is small [27].

Finally, out-of-fold predictions from all outer test folds were stored for calculation of the final results, generation of summary tables and plots. Hyperparameters chosen in the inner loop were later reused as fixed configurations for the corresponding bootstrap analyses described in the following section.

#### Bootstrapping

In this work, we use two kinds of bootstrapping. The first is intrinsic to certain models, like the Random Forest algorithm. The second is applied externally to each model and used for model evaluation. In general, bootstrapping is a resampling technique where new training sets are created by randomly resampling observations **with replacement**. These samples are called bootstrap samples. Each bootstrap sample has the same size as the original data, but includes duplicated observations while leaving others out.

As for certain models such as Random Forests or Random Survival Forests, bootstrapping is inherently applied as part of the model construction. Each decision tree is trained on a bootstrap sample drawn from the training set, with replacement. Different trees are trained on varying subsets, which introduces model diversity, reduces variance through ensemble averaging, and enables Out-of-Bag (OOB) error estimation. OOB refers to the approximately one-third of training samples left out in each bootstrap iteration. These excluded samples serve for prediction, yielding an unbiased estimation of the model’s prediction error without requiring a separate validation set. During prediction, the individual tree outputs are aggregated, commonly by averaging for regression or majority voting for classification, which also mitigates overfitting [28].

To assess model stability under limited sample size, we additionally performed an external bootstrap procedure. In each iteration, the model was retrained on a resampled dataset and evaluated on the corresponding out-of-bag (OOB) samples, i.e. , the patients not included in that resample. The aggregated bootstrap estimates characterise variability in predictive performance; in each external bootstrap, the training samples are used to fit the model. If the model itself employs internal bootstrapping (e.g. Random Forests), these internal samples are drawn from the externally resampled training set, and their own OOB estimates are computed accordingly.

External bootstrapping is applied for each of the eleven models and data subset (**V**_0_, **V**_1_, **ΔV**). However, since there are multiple pre-processing combinations that were tested during CV, bootstrapping is only applied once for each subset, using the pre-processing and hyperparameter setup that had the best *R*^2^ or Harrell’s C-index result, using a regular or survival model, respectively. This leads to three bootstrap runs per model, one for each data subset.

To balance reliability and computational cost, we used 1,520 bootstrap iterations per run. For each run, a prediction is made on the OOB samples and the appropriate metrics, corresponding to the model type, are calculated. After all runs, the mean is taken and stored as the final result for this model and data subset. These repeated evaluations further enable the stability analysis, which is described in the following section.

#### Model Stability

Model stability is the sensitivity of a machine learning algorithm to variations in the training data [29]. A model that is unstable, exhibits large changes in its performance when trained on slightly different subsets of data, whereas a stable model produces results that are more consistent. Evaluating stability complements standard performance metrics, as it is not sufficient for a model to perform well on average, but it should ideally also exhibit consistent performance across repeated runs.

In this work, we quantified model stability using the standard deviation of performance metrics across repeated cross-validation and external bootstrap runs described in Sections Cross-Validation and Bootstrapping. For basic (non-survival) models, we used standard regression metrics such as *R*^2^ and Root Mean Square Error (RMSE) [25]. Survival models were evaluated using survival-specific metrics, which are described in more detail in Section Survival Evaluation Metrics. Smaller standard deviations indicate greater stability and reliability of the model. Additionally, confidence intervals computed as described in Cross-Validation provide an estimate of the uncertainty in the mean performance across runs.

#### Statistical Significance and Model Comparison

To distinguish meaningful biological signals from noise while accounting for the extensive model selection process, we employed a two-stage statistical validation framework comprising multiplicity correction and permutation testing.

##### Correction for Selection Bias (Multiplicity)

We acknowledge that exploring multiple model configurations introduces a risk of selection bias (data snooping). For mitigation, we applied the Benjamini-Hochberg (FDR) correction across all 203 tested model pipelines (combinations of algorithms, preprocessing, and filters). Adjusted p-values (*q*-values) were calculated using the distribution of performance scores across repeated cross-validation folds (174 per model). These *q*-values, reported in Tables 3– 5 indicate stability of the selected models across data splits (FDR *<* 0.001), confirming that the identified signals are robust to the model selection process.

##### Confirmatory Permutation Testing

For confirmatory prognostic validation of the final selected models, we employed the permutation test framework described by Ojala and Garriga [30], adapted for censored survival data following Uno et al. [31]. A null distribution was generated by randomly permuting the aggregated out-of-bag (OOB) predicted risk scores 10, 000 times against the fixed survival times. The observed C-index was compared against this distribution to derive a nominal p-value testing the null hypothesis of no association.

##### Model Comparison

To determine whether the predictive performance differed significantly between **V**_0_ and **ΔV** models, we utilised a non-parametric unpaired permutation test following the principles outlined by Diettrich [32]. Since the models were trained on independent datasets, the combined predictions were shuffled 10, 000 times to generate a null distribution of the mean difference, deriving a p-value to test the hypothesis that both models capture equivalent prognostic information.

#### Feature frequencies and Feature Importance

To identify the specific immune features that drive model predictions, this study quantified both feature stability and impact. The selection frequency and predictive importance of each feature across all model configurations was computed as follows.

**Feature frequency** was computed by counting how many times a feature was present in a CV or bootstrap run, i.e. how many times a feature was not filtered out by neither the feature nor the target correlation. For each outer CV fold, the model and its features were is stored in a list. After all folds have run successfully, each feature occurrence was counted and the value was normalised for better interpretability. This process was repeated for each data subset and filter setup as well as for each model and gives us the possibility to extract the values for the best model used on a particular filter setup and data subset. The same process was followed for bootstrapping.

**Feature importance** in our case describes how much a specific feature influences a model’s prediction. The approach to measure feature importance differs depending on run type (CV or bootstrapping). Frequency and Feature importance scores are generally created for each individual filter setup and data subset per model. This results in a model performance measurement table and feature importance table for each CV and bootstrap run, respectively. To compute feature importance, we use SHAP values.

To calculate a final feature importance score for a single model using the combined information of the CV runs and bootstrap runs, both, CV and bootstrap, we trained a final model on the complete dataset *X* and trained a SHAP explainer on this model to accumulate the SHAP values for both CV and bootstrap. **Note:** an important clarification is that for this part of the analysis, we focused on which features play a more important role than others in the model’s predictive performance. Therefore, **only for the SHAP values and the resulting feature importance scores**, we trained a final model using the best parameters from the corresponding CV or bootstrap run and fitted it **to the complete dataset** *X*. We clearly distinguish this final model from the models that were used to evaluate the model’s generalisation (the predictive performance throughout the CV or bootstrap run) which was done using separate folds and hold out sets as described in the corresponding chapters. Hence, the final model is used strictly for interpretative purposes. This approach aims to ensure that the interpretability analysis benefits from the maximum amount of available data while maintaining an unbiased performance assessment.

To revise what we now have (regarding a single model since the process is generally the same for each model): A feature frequency and importance measurement for each feature and filter setup and dataset, respectively (8 filter setups and 3 datasets result in 24 CV runs and 3 bootstrap runs using only the best filter setup on each of the three data subsets). To bring this information together and create one combined feature importance ranking, the four resulting values were aggregated into a single score as follows. Firstly, we took the mean of the 8 filter setups from CV + the best bootstrap setup results for each of the four values, while analysing each data subset independently. For interpretability, we then normalised the SHAP CV and bootstrap values. The aggregated feature score was then calculated:

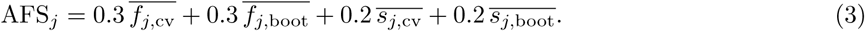

where:

AFS*_j_*= the aggregated feature score for feature *j*

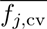= mean selection frequency of feature *j* across all cross-validation folds

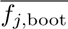 = mean selection frequency of feature *j* across all bootstrap samples

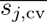 = normalised mean SHAP importance of feature *j* in cross-validation

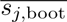 = normalised mean SHAP importance of feature *j* in bootstrapping

We acknowledge that this AFS calculation relies on fixed weights and SHAP values derived from a model fitted to the complete dataset. While this maximises information usage in a small cohort, it does not account for conditional importance among correlated predictors, meaning rankings may be influenced by the specific model structure and collinearity.

Moreover, for visual interpretation, a SHAP summary plot was created for each filter setup and data subset. Then, for interpretation, the plots that are shown down below are only from the best-performing model to ensure that the resulting feature attribution reflects the patterns learned by a robust model rather than artifacts from weaker ones that failed to generalise or detect relevant signals in the data. Hence, the plots shown in Results are only from the best model by type and data subset, showing accumulated results for all filter setups per data subset.

### Models and Approach

We evaluated two model families: regular regression models, which directly predict overall survival time and assume fully observed outcomes, and survival models, which explicitly account for right-censored time-to-event data. An overview of all evaluated regular and survival models, including their key properties, is provided in Tables 1 and 2, respectively. Hyperparameter grids are provided in the Supporting information.

**Table 1.**
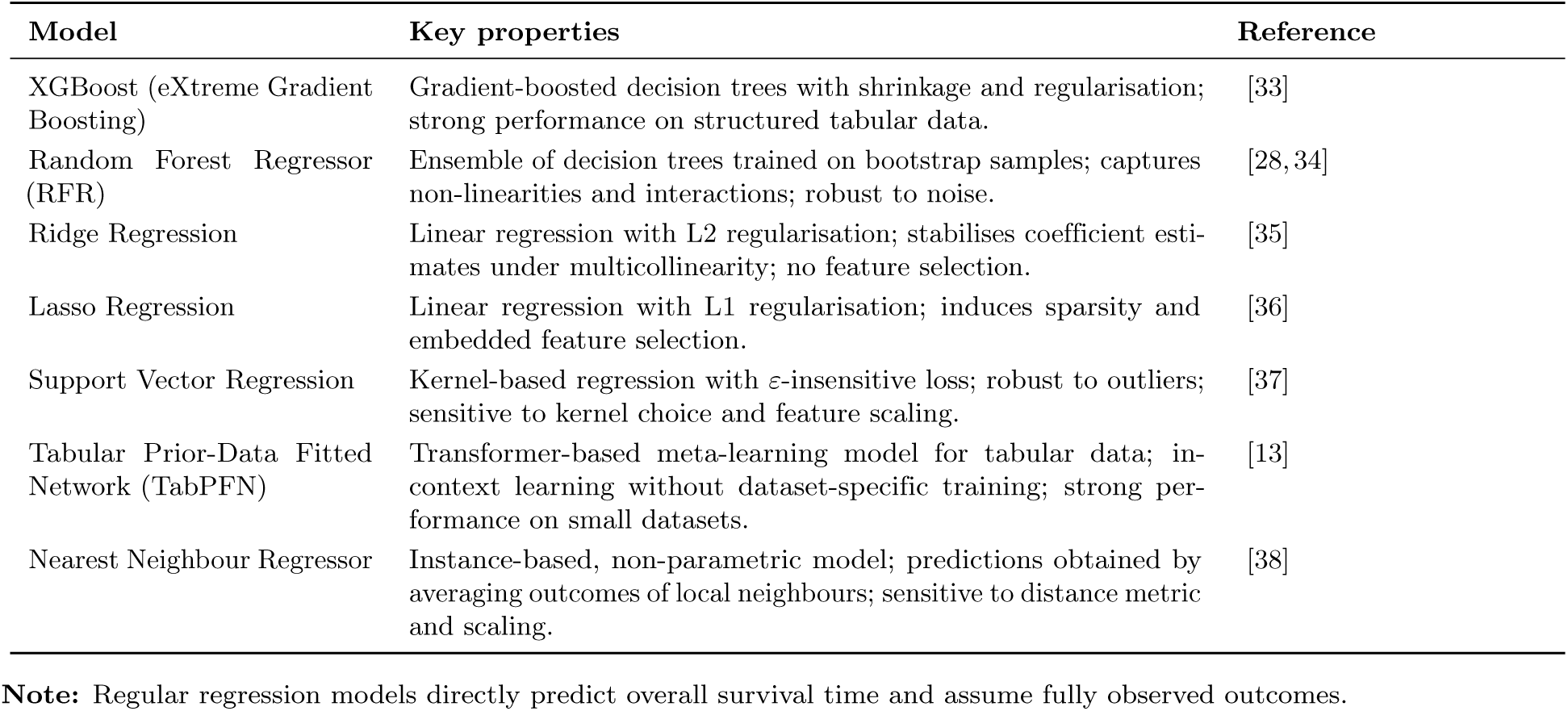
Overview of evaluated regular regression models.

| Model | Key properties | Reference |
| --- | --- | --- |
| XGBoost (eXtreme Gradient Boosting) | Gradient-boosted decision trees with shrinkage and regularisation; strong performance on structured tabular data. | [33] |
| Random Forest Regressor (RFR) | Ensemble of decision trees trained on bootstrap samples; captures non-linearities and interactions; robust to noise. | [28, 34] |
| Ridge Regression | Linear regression with L2 regularisation; stabilises coefficient estimates under multicollinearity; no feature selection. | [35] |
| Lasso Regression | Linear regression with L1 regularisation; induces sparsity and embedded feature selection. | [36] |
| Support Vector Regression | Kernel-based regression with ε-insensitive loss; robust to outliers; sensitive to kernel choice and feature scaling. | [37] |
| Tabular Prior-Data Fitted Network (TabPFN) | Transformer-based meta-learning model for tabular data; in-context learning without dataset-specific training; strong performance on small datasets. | [13] |
| Nearest Neighbour Regressor | Instance-based, non-parametric model; predictions obtained by averaging outcomes of local neighbours; sensitive to distance metric and scaling. | [38] |

**Note:** Regular regression models directly predict overall survival time and assume fully observed outcomes.

**Table 2.** Overview of evaluated survival models.

| Model | Key properties | Reference |
| --- | --- | --- |
| Random Survival Forest (RSF) | Ensemble of survival trees; non-parametric; captures non-linear effects and feature interactions; robust to model misspecification. | [39] |
| Classical Cox proportional hazards (CoxPH) | Semi-parametric survival model with log-linear hazard; assumes proportional hazards over time; interpretable hazard ratios. | [40] |
| Penalized Cox regression (CoxNet) | Cox model with L1/L2 penalties; stabilises high-dimensional fits; enables embedded feature selection. | [41] |
| Fast Survival Support Vector Machine (FastSurvivalSVM) | Ranking-based survival SVM; optimises risk ordering under right-censoring; complementary to Cox-based approaches. | [42] |

**Note:** Survival models explicitly account for right-censored time-to-event data.

### Survival Evaluation Metrics

#### C-index (Concordance Index)

measures the model’s ability to order pairs of subjects correctly by their predicted risk or survival time. It quantifies how often the model’s predictions agree with the actual ordering of observed outcomes. It is defined as:

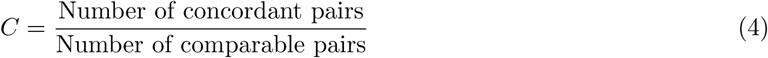

#### Concordant pair

a pair of subjects (*i, j*) where the subject with the higher predicted risk (or shorter predicted survival time) actually experiences the event before the other.

#### Comparable pair

a pair of subjects where the event times can be ordered (i.e. not both censored before either event occurs).

The C-index ranges from 0 (perfect inverse ranking) to 1 (perfect concordance), with 0.5 indicating random prediction performance. It generalises the area under the ROC curve (AUC) for censored survival data and is widely used to evaluate model discrimination in survival analysis [43].

#### IBS (Integrated Brier Score)

evaluates both calibration and discrimination of survival models over time by integrating the Brier Score across the time horizon. The Brier Score at a specific time *t* measures the squared difference between the predicted survival probability and the observed survival status. It is defined as:

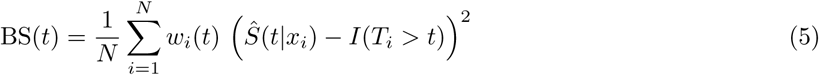

and the Integrated Brier Score is:

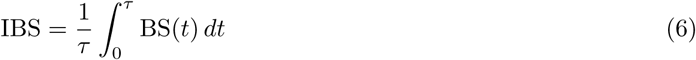

where:

*S*^^^(*t|x_i_*) is the predicted survival probability for subject *i* at time *t,*

*I*(*T_i_ > t*) is the indicator function (1 if subject *i* survived beyond time *t,* 0 otherwise),

*w_i_*(*t*) are inverse probability of censoring weights accounting for right censoring,

*τ* is the maximum follow-up time, and

*N* is the number of subjects.

The IBS ranges from 0 to 1, with lower values indicating better predictive performance. It captures both discrimination and calibration, making it a comprehensive measure of survival model accuracy [44].

### Model comparison

In small right-censored cohorts, C-index and IBS are standard endpoints for risk discrimination and calibration [43, 44]. However, cross-model comparisons are limited because *R*^2^ and C-index are not directly comparable; we therefore interpret differences as support for censoring-aware survival modelling rather than absolute superiority.

## Results

Across all analysed data subsets (**V**_0_, **V**_1_, **ΔV**), standard regression models failed to generalise (typically *R*^2^ ≤ 0); therefore, detailed performance metrics for these approaches are provided in S1 Appendix, while the main text focuses on the performance of censoring-aware survival models.

### Data Subset V**_0_**

Models were evaluated on **V**_0_ to establish baseline performance and analyse signals between feature subsets and overall survival.

**For survival models**, (Table 3), the Random Survival Forest (RSF) on **V**_0_ achieved the best overall C-index (0.656) and IBS (0.190) across all data subsets, utilising standardised data and feature-target correlation. The bootstrap IBS scores include nan values throughout all data subsets. This is due to an insufficient number of test samples, necessary for a reliable IBS score. Hence, these scores were replaced by nan values.

**Table 3.** V_0_: Best Performing Survival Models.

| Model | Standardised | Correlation | Target Corr | Run Type | C-Index | C 95 % CI | IBS | IBS 95 % CI | FDR $q$-value |
| --- | --- | --- | --- | --- | --- | --- | --- | --- | --- |
| RSF | True | False | True | CV | **0.656** | **[0.636, 0.677]** | **0.190** | **[0.181, 0.199]** | < **0.001\*** |
| RSF | False | False | True | CV | 0.654 | [0.634, 0.675] | 0.192 | [0.183, 0.200] | < **0.001\*** |
| RSF | True | True | True | CV | 0.618 | [0.597, 0.638] | 0.201 | [0.192, 0.209] | < **0.001\*** |
| RSF | True | False | True | BOOT | 0.623 | [0.355, 0.857] | nan | [nan, nan] | < **0.001\*** |

**Note:** Top three CV runs sorted by C-index. The BOOT run corresponds to the best CV run. IBS BOOT values are nan due to insufficient test samples. **Abbreviations:** RSF (Random Survival Forest). **FDR $q$ -value:** Benjamini-Hochberg adjusted p-value correcting for multiplicity; $q \leq 0.05$ indicates statistical significance.

### Data Subset V**_1_**

**V**_1_ was included to determine if absolute feature values after two cycles of ICT correlate with OS.

**Survival Models** (Table 4) on **V**_1_ had the lowest predictive power overall. The best RSF setup reached a C-index of only 0.544.

**Table 4.** V_1_: Best Performing Survival Models.

| Model | Standardised | Correlation | Target Corr | Run Type | C-Index | C 95 % CI | IBS | IBS 95 % CI | FDR $q$-value |
| --- | --- | --- | --- | --- | --- | --- | --- | --- | --- |
| RSF | True | True | False | CV | 0.544 | [0.520, 0.568] | 0.214 | [0.206, 0.222] | 0.307 |
| RSF | False | False | False | CV | 0.541 | [0.517, 0.566] | 0.218 | [0.210, 0.225] | 0.307 |
| RSF | True | False | False | CV | 0.534 | [0.512, 0.556] | 0.215 | [0.207, 0.223] | 0.307 |
| RSF | True | True | False | BOOT | 0.541 | [0.200, 0.833] | nan | [nan, nan] | 0.307 |

**Note:** Top three CV runs sorted by C-index. BOOT corresponds to the best CV.

### Data Subset ΔV

**ΔV** assesses whether dynamic changes in immune cell composition correlate with overall survival.

**Survival models** (Table 5) reached results comparable to **V**_0_, with the RSF achieving a C-index of 0.649. However, the bootstrap CI (0.285 to 1.000) was significantly wider than that of **V**_0_, indicating lower prediction confidence.

**Table 5.** ΔV: Best Performing Survival Models.

| Model | Standardised | Correlation | Target Corr | Run Type | C-Index | C 95 % CI | IBS | IBS 95 % CI | FDR $q$-value |
| --- | --- | --- | --- | --- | --- | --- | --- | --- | --- |
| RSF | False | False | True | CV | 0.649 | [0.620, 0.678] | 0.209 | [0.197, 0.220] | < **0.001\*** |
| RSF | True | False | True | CV | 0.643 | [0.616, 0.671] | 0.207 | [0.196, 0.217] | < **0.001\*** |
| RSF | False | True | True | CV | 0.634 | [0.605, 0.663] | 0.211 | [0.201, 0.222] | < **0.001\*** |
| RSF | False | False | True | BOOT | 0.640 | [0.285, 1.000] | nan | [nan, nan] | < **0.001\*** |

**Note:** Top three CV runs sorted by C-index. BOOT corresponds to the best CV.

### Statistical Signal Verification

Evaluation of model stability using Benjamini-Hochberg correction confirmed robust performance across model configurations (FDR *q <* 0.001, see Table 3 and Table 5). Confirmatory permutation testing subsequently validated the presence of prognostic signal in **V**_0_ (*p* = 0.015) and **ΔV** (*p* = 0.022), whereas the **V**_1_ model failed to show significance (*p* = 0.445), confirming a lack of significant global discrimination in absolute post-treatment values under our evaluation.

#### Model Comparison

Direct comparison of the predictive models confirmed that **V**_0_ and **ΔV** models were statistically distinguishable from **V**_1_, and from each other (Unpaired Permutation Test difference = 2.8%, *p <* 0.001).

Critically, feature importance analysis shows that the models prioritise distinct sets of features (See 6, 7,4,5), countering the hypothesis that the **ΔV** model is primarily a variance-reduced version of the baseline. This divergence in feature utilisation confirms that the **ΔV** model captures independent prognostic information. Boxplots of the results are available in S1 Fig.

### Feature Rankings V**_0_** and ΔV

Tables 6 and 7 present the top 20 features for the best performing RSF models, ranked by their accumulated feature score (AFS*_j_*). This score aggregates frequency and SHAP importance across cross-validation and bootstrap runs. The full list of features is available in S1 Table.

**Table 6.**
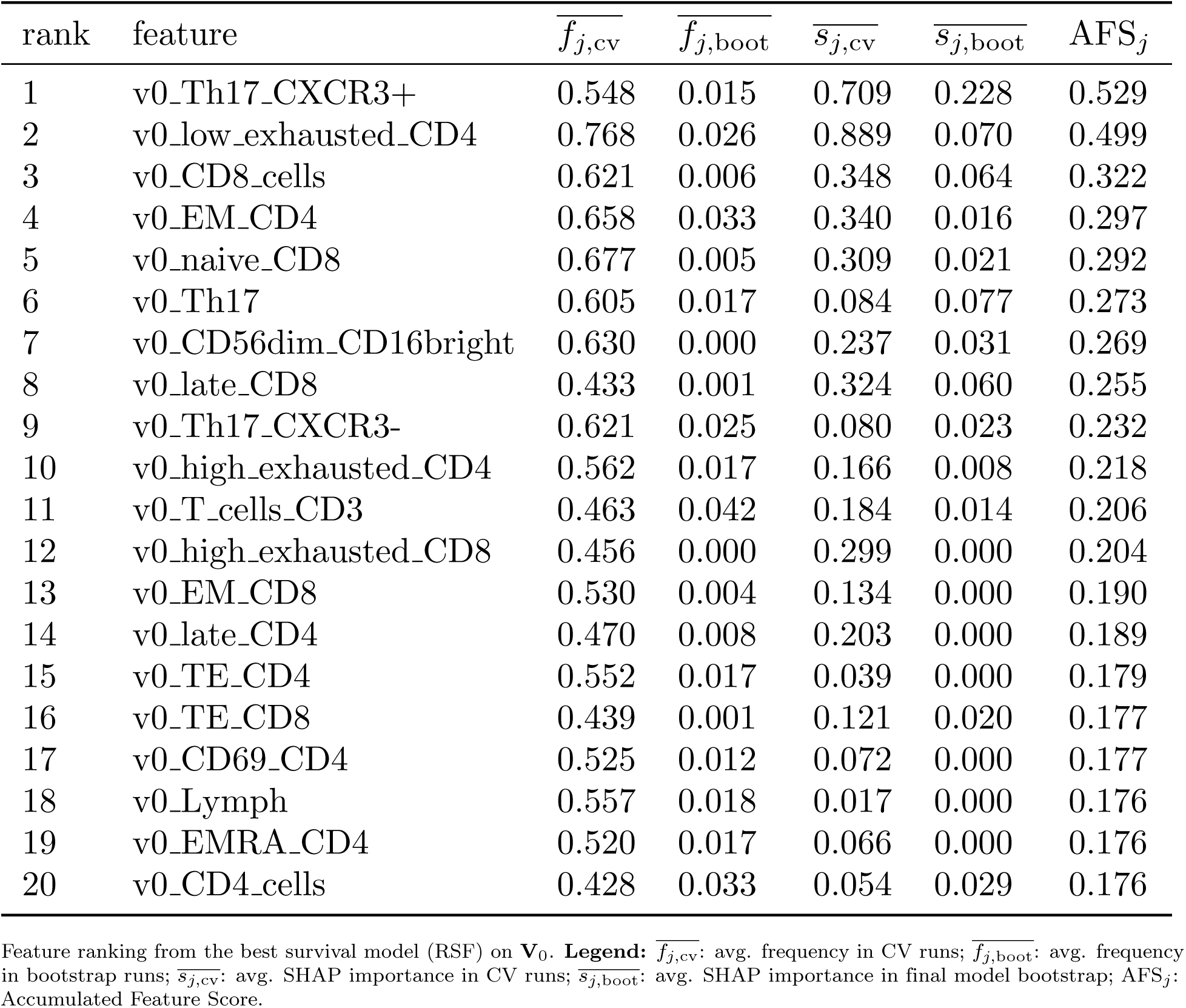
Feature Ranking Top 20 Best Survival Model V_0_.

**Table 7.** Feature Ranking Top 20 Best Survival Model ΔV.

| rank | feature | $\overline{f_{j,cv}}$ | $\overline{f_{j,boot}}$ | $\overline{s_{j,cv}}$ | $\overline{s_{j,boot}}$ | AFS<sub>j</sub> |
| --- | --- | --- | --- | --- | --- | --- |
| 1 | delta_naive_Treg_CD4 | 0.718 | 0.003 | 1.023 | 0.184 | 0.616 |
| 2 | delta_Th17_CXCR3+ | 0.777 | 0.014 | 0.181 | 0.084 | 0.364 |
| 3 | delta_non_class_switched_memory_B | 0.641 | 0.026 | 0.463 | 0.041 | 0.335 |
| 4 | delta_CD4_CD8_DP | 0.829 | 0.000 | 0.251 | 0.032 | 0.332 |
| 5 | delta_Th17 | 0.722 | 0.009 | 0.255 | 0.054 | 0.328 |
| 6 | delta_Th17_CXCR3- | 0.573 | 0.010 | 0.254 | 0.077 | 0.308 |
| 7 | delta_Th1 | 0.713 | 0.002 | 0.411 | 0.000 | 0.295 |
| 8 | delta_naive_CD8 | 0.709 | 0.003 | 0.241 | 0.026 | 0.289 |
| 9 | delta_EMRA_CD4 | 0.610 | 0.018 | 0.231 | 0.031 | 0.268 |
| 10 | delta_interm_CD4 | 0.798 | 0.002 | 0.043 | 0.000 | 0.248 |
| 11 | delta_CD56dim_CD16bright | 0.733 | 0.000 | 0.077 | 0.000 | 0.235 |
| 12 | delta_class_switched_memory_B | 0.754 | 0.005 | 0.029 | 0.000 | 0.233 |
| 13 | delta_Lymph | 0.572 | 0.017 | 0.168 | 0.021 | 0.233 |
| 14 | delta_low_exhausted_CD8 | 0.699 | 0.000 | 0.029 | 0.000 | 0.215 |
| 15 | delta_CD8_cells | 0.457 | 0.003 | 0.052 | 0.034 | 0.185 |
| 16 | delta_T_cells_CD3 | 0.490 | 0.034 | 0.062 | 0.014 | 0.184 |
| 17 | gender | 0.547 | 0.011 | 0.000 | 0.000 | 0.167 |
| 18 | delta_naive_Treg_CD8 | 0.483 | 0.000 | 0.088 | 0.000 | 0.162 |
| 19 | delta_Treg_CD4_CD127low | 0.520 | 0.004 | 0.000 | 0.000 | 0.157 |
| 20 | delta_TE_CD4 | 0.497 | 0.007 | 0.016 | 0.000 | 0.154 |

Feature ranking from the best survival model (RSF) on $\Delta\mathbf{V}$ . **Legend:** See Table 6 for variable definitions.

### SHAP Beeswarm Plot V_0_ and ΔV

Fig 4 and Fig 5 illustrate the SHAP beeswarm plots for the best RSF models, detailing the top 15 features and the directionality of their impact on survival risk.

**Fig 4.**
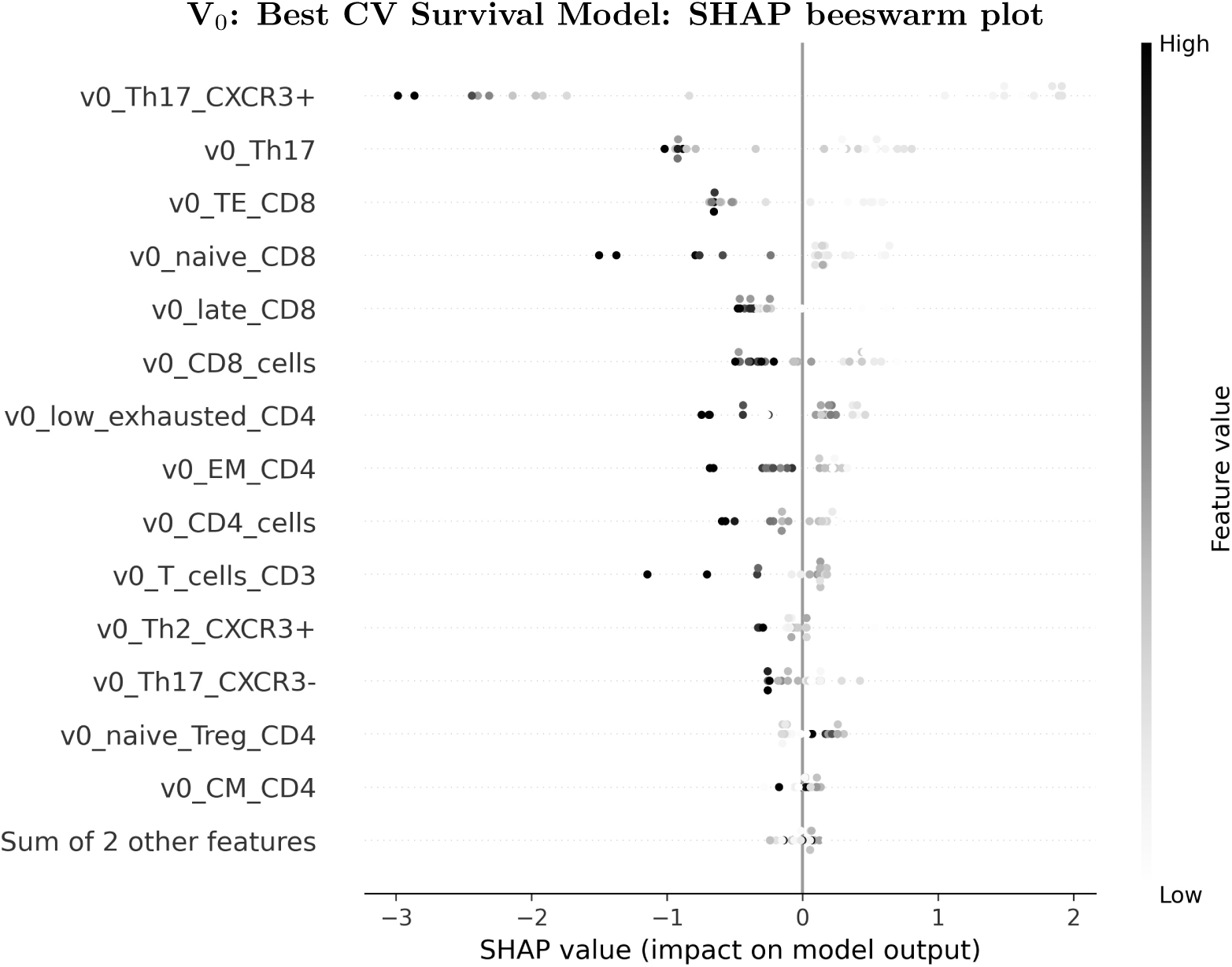
**V_0_**: SHAP beeswarm plot for the best survival model (CV). The x-axis represents the SHAP value; positive values indicate a higher predicted risk (i.e. shorter survival), while negative values indicate a lower risk (i.e. longer survival). Grey-scales represent feature values (i.e. black = high and white = low.)

**Fig 5.**
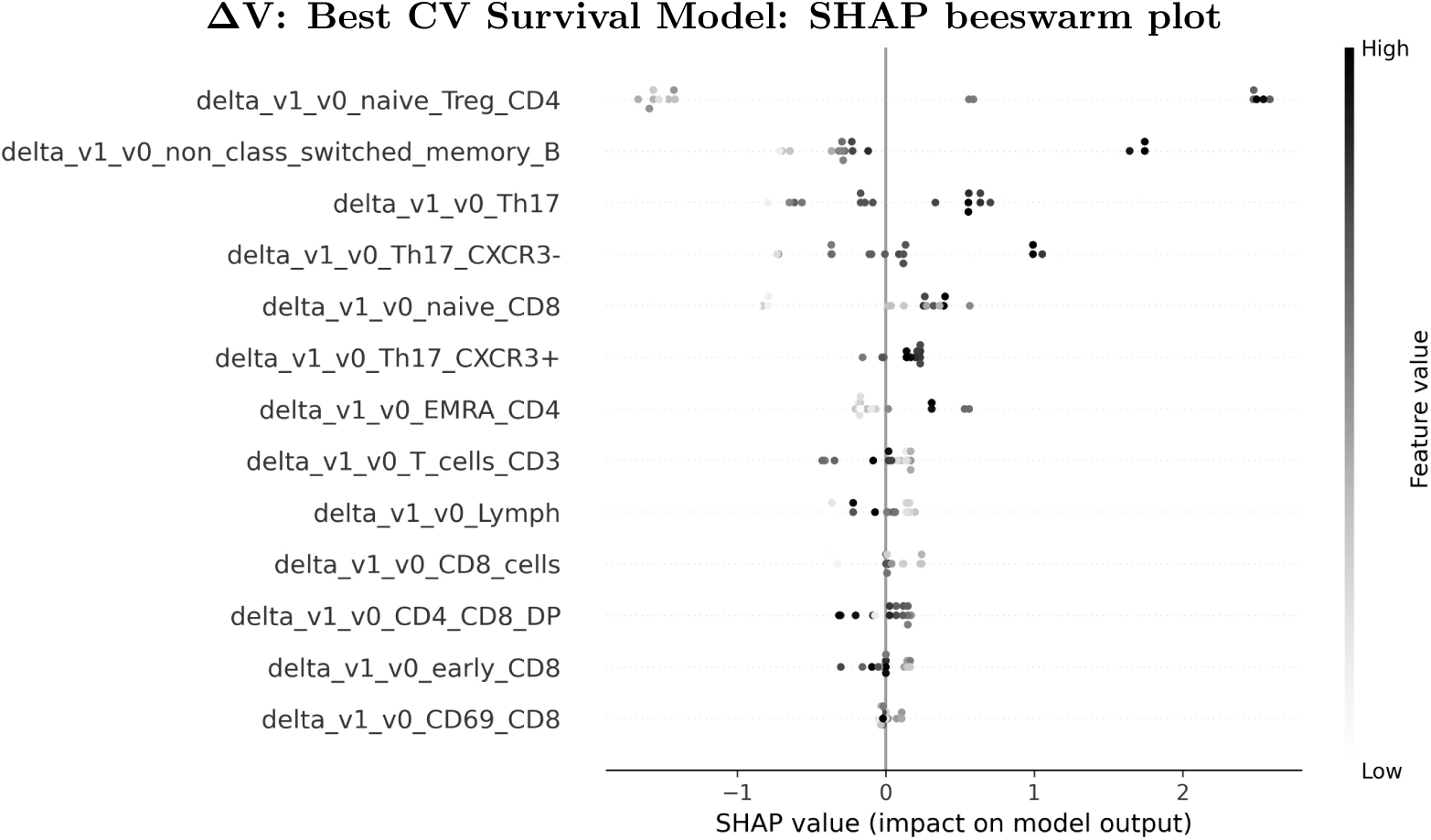
**ΔV**: SHAP beeswarm plot for the best survival model (CV). The x-axis represents the SHAP value, as described in Fig. 4.

## Discussion

In this study we applied a comprehensive machine learning framework to evaluate predictive value of peripheral lymphocyte subsets in stage IV SCLC patients treated with immunochemotherapy (ICT). Given the constraints of a limited cohort size (*n* = 32), the primary challenge was distinguishing true biological signals from the high variance inherent in small-scale clinical data. To address this, we employed a methodological approach specifically designed to stress-test the stability of our findings before drawing biological conclusions.

### Methodological Grounding: Validation and Stability

To ensure that our results were not artifacts of overfitting, we utilised a rigorous validation pipeline. We implemented a nested cross-validation (CV) scheme, separating hyperparameter tuning (inner loop) from performance evaluation (outer loop). This ensured that the model was never evaluated on data it had seen during optimisation. Furthermore, to assess the reliability of these predictions, we applied bootstrapping (1, 520 iterations), where models were retrained on resampled datasets and evaluated on out-of-bag (OOB) samples.

It is within this conservative framework that we evaluated our models. Standard regression algorithms like Ridge Regression and XGBoost failed to generalise, yielding negative *R*^2^ scores across all subsets (**V**_0_, **V**_1_, and **ΔV**). This indicates that the relationship between immune subsets and survival time is likely non-linear and heavily influenced by censoring, which standard regression loss functions fail to accommodate. However, survival models and specifically Random Survival Forests (RSF) successfully extracted predictive patterns and achieved concordance indices (C-index) of approximately 0.66 for baseline (**V**_0_) and 0.65 for delta (**ΔV**) datasets.

### Statistical Validation of the Signal

Given the small dataset size, a C-index of 0.66, while promising, carries a risk of being a stochastic anomaly. To mitigate this, we derived our performance metrics (*R*^2^ and RMSE for regular models and C-index and

Integrated Brier Score (IBS) for survival models) by averaging them across all 174 nested cross-validation folds, yielding a conservative and more realistic estimate of model stability quantified by 95% confidence intervals. To further ground this result and confirm the presence of a true signal, we relied on the permutation test framework described in Statistical Significance and Model Comparison.

#### Robustness of Performance Estimates

the confidence intervals (CI) obtained during our validation are used to evaluate the robustness and performance estimates of the models. In the nested CV, the 95% CIs for the C-index were notably narrow (e.g. **V**_0_ 0.638 0.680). This narrowness reflects the high number of repetitions used to stabilise the performance estimate itself which presents a convergence towards the mean values of the metric if models performed well. However, the external bootstrap CIs were significantly wider and crossed the 0.5 threshold (e.g. **V**_0_ 0.333 0.857). This discrepancy shows that while our estimation procedure is robust (narrow CV CI), the underlying model remains sensitive to specific data splits due to the small sample size (wide bootstrap CI).

#### Calibration (IBS)

Beyond ranking patients correctly (C-index), we evaluated calibration using the Integrated Brier Score (IBS). The best **V**_0_ model achieved an IBS of 0.191 (95% CI: 0.182−0.200). Since lower IBS values indicate better calibration, this score suggests that the model’s predictive survival probabilities are reasonably accurate and not merely effective at ranking risk.

#### Signal Verification

As reported in the Results (Statistical Signal Verification), permutation testing confirmed statistically significant signals for both the baseline **V**_0_ (*p* = 0.015) and for the models evaluated on **ΔV** (*p* = 0.022). In contrast, the models trained on **V**_1_ – data of patients after two cycles of ICT – yielded a non-significant p-value of 0.445. This distinct lack of significance in **V**_1_ serves as a validation of our statistical framework, demonstrating that the test correctly discriminates between predictive signal (baseline and delta) and random noise (absolute values after 2 cycles of ICT).

#### Model Comparison

The unpaired permutation test confirmed that the predictions of the baseline **V**_0_ and **ΔV** models were significantly different (*p* = 0.0001). This implies that the prognostic information contained in the pre-treatment immune state is statistically distinct from the information contained in the early therapy-induced immune shifts.

### Biological Implications: V**_0_** and ΔV

Our analysis provided statistical evidence supporting the first hypothesis (*H*_1_), demonstrating that the baseline composition of peripheral blood prior to ICT is significantly associated with survival. The predictive strength of the model, however, remains modest (C-index approx. 0.66), underscoring the necessity for caution when interpreting these results, especially given the small data size. Hence, we interpret this finding as a **proof of concept:** that an immunological signal exists, even if it is inherently weak and noisy in this small cohort.

Importantly, regarding the second hypothesis (*H*_2_), we observed a high contrast between absolute values after two cycles of ICT (**V**_1_) and dynamic changes (**ΔV**). The **V**_1_ dataset yielded no significant signal (*p* = 0.445), whereas the **ΔV** dataset was significant (*p* = 0.022). This supports the conclusion that for SCLC patients, the **direction and magnitude** of the immune shift are more clinically relevant than the absolute cell counts achieved after two cycles of ICT. Notably, this predictive signal was detected in the **ΔV** subset (*n* = 16) despite it having a smaller sample size than the non-significant **V**_1_ subset (*n* = 20), further reinforcing the potency of the dynamic signal.

The Aggregated Feature Score (AFS), which synthesises feature selection frequency and SHAP importance, used in the RSF model for **ΔV**, confirmed the predictive value of delta Th17 (i.e., Th17 cell reduction), but notably identified Delta Naive Regulatory T cells ((Tregs), `delta_naive_Treg_CD4`) as the highest-ranked feature. This high ranking, derived from both its stability across cross-validation and its high contribution to the model output, suggests a complex interplay where the modulation of Tregs, which mediate immunosuppression, alongside Th17 normalisation to healthy donor levels [8], plays a pivotal role in the early response to ICT. Tregs are known to dampen immune responses and prevent excessive inflammation [8]. Hence, their dynamic fluctuation likely reflects the immune system’s attempt to recalibrate between autoimmune regulation and anti-tumour activity under the influence of PD-L1 blockade.

Moreover, the identification of Naive Regulatory T cells and Memory B cells as key features aligns with emerging evidence in cancer immunotherapy. While Tregs are generally associated with immunosuppression, recent studies suggest that the dynamic replenishment of the naive Treg pool may reflect a systemic reset of immune tolerance mechanisms required for checkpoint inhibition efficacy [45]. Similarly, B cells are increasingly recognised as critical partners in the anti-tumor immune response, particularly through the formation of tertiary lymphoid structures (TLS) which support T cell activation [46]. The prioritisation of these features by the RSF model–distinct from the Th17 signal alone–suggests that the machine learning pipeline has successfully captured a multi-lineage immune coordination necessary for effective ICT response.

#### Comparison with Prior Findings

Our results provide an expansion of the findings reported by Schmälter et al. [8]. A key distinction lies in the baseline (**V**_0_) analysis. While the prior study – relying on standard univariate statistical tests – did not observe a significant prognostic value in pretreatment intervals, our machine learning approach detected a statistically significant signal (*p* = 0.015). This suggests that the relationship between baseline immune subsets and survival is likely non-linear and driven by multivariate interactions, patterns that classical linear regression or log-rank tests may fail to capture in small, high-variance cohorts.

Regarding specific features, both studies converge on the importance of Th17 cells. Schmälter et al. [8] identified a reduction in Th17 as a potential survival marker, and our analysis consistently ranked `delta_Th17_CXCR3+` and `delta_Th17` among the top predictors. However, the prior study highlighted CD4 Central Memory (CM), Early CD4, and CD8 CM subsets as significant. In contrast, our Aggregated Feature Score (AFS) did not prioritise these features. This discrepancy likely arises from methodological differences:

#### Feature Filtering

Our pipeline utilised a correlation-based filter during preprocessing. If these memory subsets were highly correlated with other dominant features (redundancy) or lacked a strong individual correlation with the target in the training folds, they may have been filtered out or de-prioritised during the permutation importance calculations.

#### Metric Definition (SHAP vs. Log-Rank)

The prior study utilised Kaplan-Meier estimators and log-rank tests. Different approaches of these tests weight time-points differently, potentially emphasising early vs. late events. In contrast, our SHAP values represent the marginal contribution of a feature to the model’s prediction across the entire dataset. A feature might be statistically significant in a univariate curve comparison but have lower predictive utility in a multivariate forest when other stronger features are available.

#### Robustness of Ranking (AFS)

The feature lists presented in this paper rely on the Aggregated Feature Score (AFS). Unlike a single p-value, the AFS synthesises feature selection frequency across cross-validation and bootstrapping with SHAP importance. This ensures that the reported top features are not merely artifacts of a single successful run but are persistently important across hundreds of resampled data splits. Since CV and Bootstrapping were used, we cannot present a single feature set per run, but the accumulation of features that regularly passed the correlation filters and that valued as important by SHAP.

Finally, the success of the **ΔV** models, dominated by the Random Survival Forest, highlights the utility of dynamic changes. In small cohorts, high inter-patient variability (background noise) makes it difficult to detect group differences. By calculating the delta, we effectively normalise the data to each patient’s own baseline, removing static noise and isolating the biologically relevant signal.

### Limitations

#### Sample Size and Selection Bias

The study’s primary limitation is the sample size(*n* = 32). Rapid attrition due to disease progression (*<* 10 samples after **V**_2_) restricted post-treatment analysis to two cycles after ICT (**V**_1_), meaning the observed lack of signal at **V**_1_ cannot be generalised to later timepoints. Furthermore, complete-case comparisons (**V**_0_ (*n* = 24), **V**_1_ (*n* = 20), and **ΔV** (*n* = 16)) may be subject to informative missingness; this risks inflating **ΔV** performance through survivor bias while depressing **V**_1_ estimates. Given the resulting sparsity and wide bootstrap confidence intervals, these findings represent single-centre proof of concept indicators requiring validation in larger, multi-centre cohorts.

#### Biological Scope and Compartment

Both this analysis and the pilot study evaluated peripheral blood lymphocytes in isolation. While offering a non-invasive liquid biopsy, peripheral blood is a systemic compartment influenced by physiological factors beyond the malignancy, distinct from the tumour microenvironment (TME), which is the direct site of immune evasion and has been the focus of extensive research [47–50], where immune evasion directly occurs. The absence of tumour-intrinsic data likely contributes to the unexplained variance in our models.

#### Confounding Factors

Analyses lacked adjustment for clinical confounders (e.g. corticosteroids, antibiotics, ECOG status, tumour burden). While the **ΔV** approach partially mitigates fixed confounding by normalising to patient baselines, specific feature importances should be interpreted as candidate signals requiring multivariate validation in larger cohorts.

#### Measurement and Reproducibility

Flow cytometry subset definitions and gating strategies strictly adhered to protocols previously validated by Schmälter et al. [8]. As a secondary computational analysis, this study relies on the quality control established in that primary publication and did not perform separate inter-operator reproducibility assessments or additional FMO quantification.

#### Feature Ranking Stability

The AFS utilised fixed weights and whole-dataset SHAP values without correcting for the conditional importance among correlated predictors. Consequently, rankings may be influenced by collinearity and model selection; identified features should therefore be viewed as candidate components pending robustness checks in independent datasets.

#### Computational Constraints

Due to computational constraints, permutation testing relied on permuting predictions of optimised models rather than full retraining. We mitigated the associated risk of selection bias by applying Benjamini-Hochberg FDR correction across cross-validation folds, ensuring that only models with high structural stability were evaluated for significance.

#### Methodological Comparability

Finally, we acknowledge that the direct comparison between regression and survival models is constrained by the use of differing metrics, necessitating an interpretation focused on the utility of censoring-aware modelling rather than direct equivalence.

## Conclusion

This paper successfully employed a rigorous machine learning framework to validate and expand on the potential immunological biomarkers for SCLC identified by Schmälter et al. [8]. By utilising nested cross-validation and permutation testing, we demonstrated that Random Survival Forests can extract statistically significant prognostic signals from small, high-dimensional immunological datasets, where standard regression methods fail.

Our results lead to three main conclusions:

1. **Robust Validation of Weak Signals:** Through extensive bootstrapping and permutation testing, we confirmed that the signals detected in baseline **V**_0_ and dynamic **ΔV** immune profiles are statistically significant (*p <* 0.05) and not artifacts of random chance, despite the limited sample size.
2. **Dynamic changes over Absolute States:** In this proof of concept cohort, early dynamic shifts in the balance between regulatory and effector immune arms are associated with prognosis, with significance confirmed after multiplicity-adjusted testing, contrasting with the lack of signal in absolute counts after two cycles of ICT (**V**_1_). Given attrition and complete-case pairing, this comparative finding requires confirmation with missing-data-aware analyses and external validation, but suggests that the trajectory of the immune response may be a superior early biomarker compared to static measurements.
3. **Expanded Immunological Signature:** Our multivariate analysis validated Th17 cell reduction and identified Naive Regulatory T cells and Memory B cells as candidate components of a broader signature. This suggests that an effective response to ICT involves a systematic recalibration of both the regulatory and effector arms of the immune system, though the precise ranking of these features warrants confirmation in covariate-adjusted analyses.

### Future Directions

These findings support the viability of liquid biopsy immune profiling in SCLC. Future work should focus on validating these specific dynamic signatures in larger cohorts and aim for a multimodal integration of data. As previous studies have demonstrated that circulating lymphocytes can reflect the local immune response [51], combining these systemic immune profiles with tumour-intrinsic information (such as TMB or PD-L1 status) could effectively bridge the gap between systemic noise and tumour-specific signals.

Finally, this paper highlights the relevance of a systems immunology approach. While the biological hypothesis provided the foundation, the application of machine learning enabled the detection of non-linear signals that traditional statistical methods missed. Moving forward, the integration of clinical expertise with advanced computational methods represents a promising and important direction to address the prognostic complexity of biomedical data.

## Supporting information

Regular Model Performance

Detailed Pre-Processing

Boxplots of Cross-Validation Results

Full Feature Ranking Lists

Model Hyperparameter Grids

## Data Availability

All data produced in the present study are available upon reasonable request to the authors

## Supporting information

**S1 Appendix. Regular Model Performance.** Includes the performance metrics (Tables A-C) for regular regression models evaluated on datasets **V**_0_, **V**_1_, and **ΔV**.

**S2 Appendix. Detailed Pre-Processing** Includes the detailed steps used for missing value handling, feature filtering, categorical encoding, and standardisation.

**S1 Fig. Boxplots of Cross-Validation Results.** Distribution of performance metrics (*R*^2^ and C-index) across all 174 CV folds for the top-performing regular and survival models.

**S1 Table. Full Feature Ranking Lists.** Complete tables of aggregated feature scores (frequency and SHAP) for the best survival models on **V**_0_ and **ΔV**.

**S2 Table. Model Hyperparameter Grids.** Detailed search spaces used for hyperparameter optimisation of all evaluated algorithms (Tables D-N).

**S3 Table. Data set used in this study.** The dataset restricted to the variables analysed in this study.

## Acknowledgments

We thank Dr. Torsten Straßer for his valuable advice regarding the statistical methodology. We gratefully acknowledge the patients and their families for their participation in this study.

