## Supplementary material for "AURORA: Analysing and understanding responses to oncological regimens with artificial intelligence": Regular Model Performance

Regular Model Results

### Table A: v0 - Top Models Performance

Regular Models

| Model | Dataset | Standardized | Correlation | Target Corr | Run Type | R2 | R2 95% CI | RMSE | RMSE 95% CI | FDR P-Val |
| --- | --- | --- | --- | --- | --- | --- | --- | --- | --- | --- |
| Ridge | v0 | False | False | False | CV | 0.029 | [-0.101, 0.159] | 158.103 | [151.570, 164.637] | 1.000 |
| XGBRegressor | v0 | False | False | False | CV | 0.027 | [-0.105, 0.159] | 165.085 | [157.689, 172.481] | 1.000 |
| RandomForestRegressor | v0 | True | False | True | CV | -0.042 | [-0.171, 0.088] | 170.499 | [163.143, 177.855] | 1.000 |
| Ridge | v0 | False | False | False | BOOT | -0.018 | [-2.510, 0.807] | 158.081 | [86.354, 262.165] | 1.000 |

**Note**: Top three CV runs sorted by R2. The BOOT run corresponds to the best CV run.

**Filters**: Standardised (feature scaling), Correlation (feature correlation filter), Target Corr (feature-target correlation filter).

**FDR q-value**: Benjamini-Hochberg adjusted p-value correcting for multiplicity; q ¡ 0.05 indicates statistical significance.

### Table B: v1 - Top Models Performance

Regular Models

| Model | Dataset | Standardized | Correlation | Target Corr | Run Type | R2 | R2 95% CI | RMSE | RMSE 95% CI | FDR P-Val |
| --- | --- | --- | --- | --- | --- | --- | --- | --- | --- | --- |
| SVR | v1 | False | False | False | CV | -0.500 | [-0.644, -0.356] | 202.478 | [193.449, 211.506] | 1.000 |
| XGBRegressor | v1 | True | False | False | CV | -0.589 | [-0.761, -0.417] | 206.596 | [197.387, 215.805] | 1.000 |
| XGBRegressor | v1 | True | True | False | CV | -0.594 | [-0.768, -0.421] | 206.255 | [197.255, 215.256] | 1.000 |
| SVR | v1 | False | False | False | BOOT | -3.717 | [-3.835, 0.054] | 204.786 | [97.934, 322.570] | 1.000 |

**Note**: Top three CV runs sorted by R2. BOOT corresponds to the best CV.
**Abbreviations**: SVR (Support Vector Regressor).

### Table C: Delta_v1v0 - Top Models Performance

Regular Models

| Model | Dataset | Standardized | Correlation | Target Corr | Run Type | R2 | R2 95% CI | RMSE | RMSE 95% CI | FDR P-Val |
| --- | --- | --- | --- | --- | --- | --- | --- | --- | --- | --- |
| TabPFNRegressor | Delta_v1v0 | True | False | True | CV | **0.043** | **[-0.121,0.206]** | **134.092** | **[127.496,140.687]** | 1.000 |
| TabPFNRegressor | Delta_v1v0 | False | False | True | CV | 0.033 | [-0.110, 0.175] | 135.033 | [128.203, 141.863] | 1.000 |
| TabPFNRegressor | Delta_v1v0 | True | False | False | CV | -0.002 | [-0.176, 0.171] | 137.269 | [130.428, 144.111] | 1.000 |
| TabPFNRegressor | Delta_v1v0 | True | False | True | BOOT | -0.261 | [-3.925, 0.833] | 137.876 | [45.116, 262.139] | 1.000 |

**Note**: Top three CV runs sorted by R2. BOOT corresponds to the best CV.
