## Supplementary material for "AURORA: Analysing and understanding responses to oncological regimens with artificial intelligence": Detailed Pre-Processing

### Missing Value Handling

Missing values occured across therapy cycles. Two strategies were tested:

1. KNN Imputation using scikit-learn’s KNNImputer^11^

2. Domain-informed imputation based on expert knowledge

### Dimensionality Reduction / Feature Filtering

Three filtering strategies were applied to reduce redundancy and improve model convergence:

1. Variance filter – features with variance Vcolumn < 0.1 were removed. No columns met this criterion, so this filter did not reduce features in practice.

2. Feature correlation filter – Spearman correlation was used to identify highly correlated features. Features exceeding a correlation threshold of 0.9 were removed. For CoxNet, CoxPH, and FastSurvivalSVM (see Models and Approach), a stricter threshold of 0.7 was applied.

3. Target correlation filter – Spearman correlation between each feature and the target was computed; features with p > 0.05 were removed.

All filters were tested in combination, switching each filter on or off permutatively to determine the best configuration for each model and data subset. Since the EDA indicated non-normality and outliers in the data, Spearman correlation was used instead of Pearson. Spearman’s rank-based approach is less sensitive to outliers additionally when normality of the data cannot be assumed and better captures monotonic relationships that are not necessarily linear [42, 43].

### Categorical Encoding and Standardisation

1. Categorical features (e.g. gender) were one-hot encoded.

2. Continuous features were standardised to zero mean and unit variance using scikit-learn’s StandardScaler^12^.

All pre-processing steps were applied within each training fold and bootstrap sample, respectively to avoid data leakage.

^11^scikit-learn version 1.7.1. https://scikit-learn.org/stable/modules/generated/sklearn.impute.KNNImputer.html, accessed 6 November 2025

^12^scikit learn’s StandardScaler https://scikit-learn.org/0.15/modules/generated/sklearn.preprocessing.StandardScaler.html, accessed 6 November 2025

42. Bishara AJ, Hittner JB. Reducing Bias and Error in the Correlation Coefficient Due to Nonnormality. Educational and Psychological Measurement. 2015;75(5):785–804. doi:10.1177/0013164414557639.

43. A Comparison of Monotonic Correlation Measures with Outliers. ResearchGate. 2025;.
