## Supplementary figures and images for "AURORA: Analysing and understanding responses to oncological regimens with artificial intelligence"

### Boxplots of Cross-Validation Results

#### v0 — Regular Models (Top 3 by $R^2$)

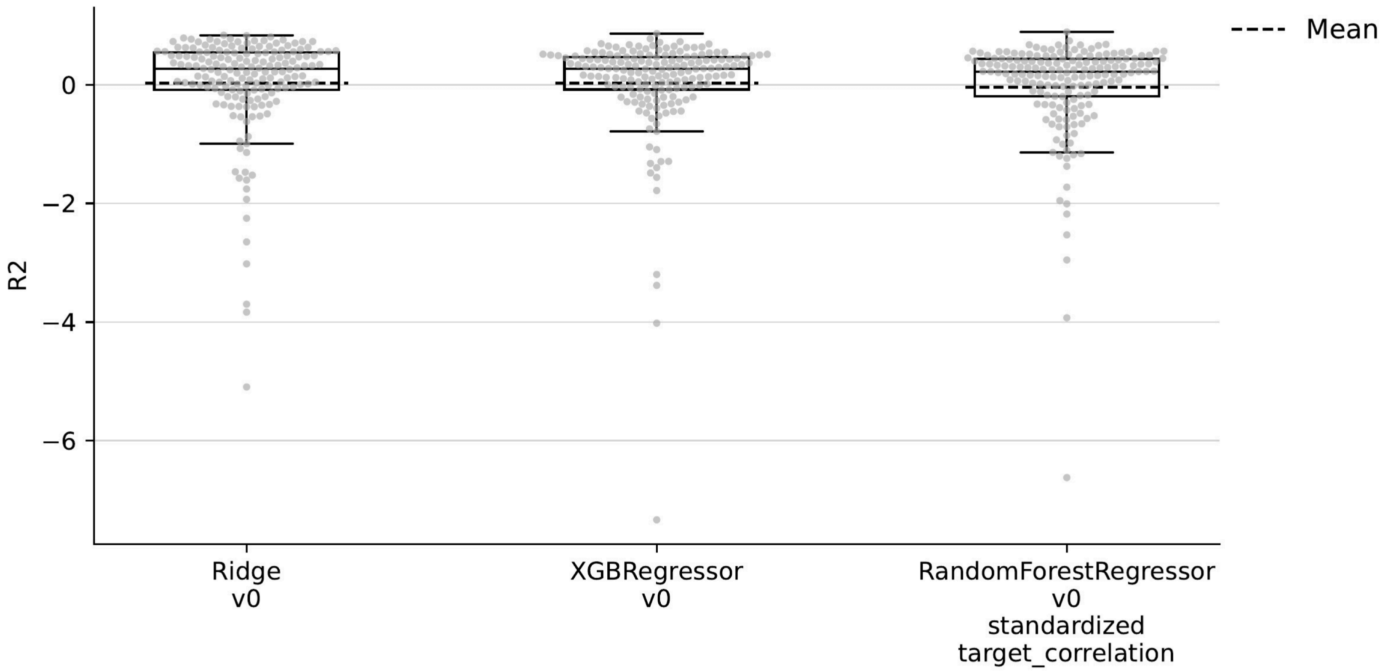

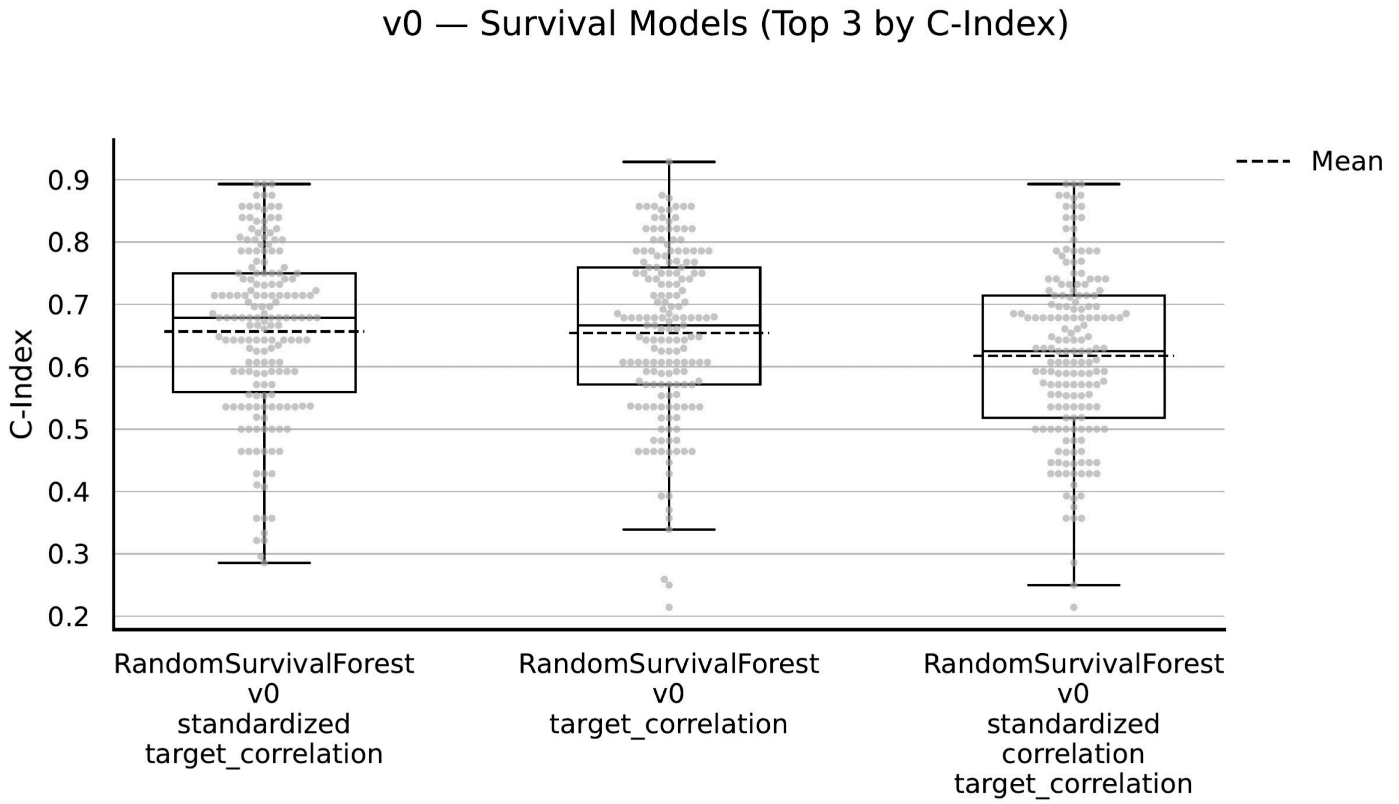

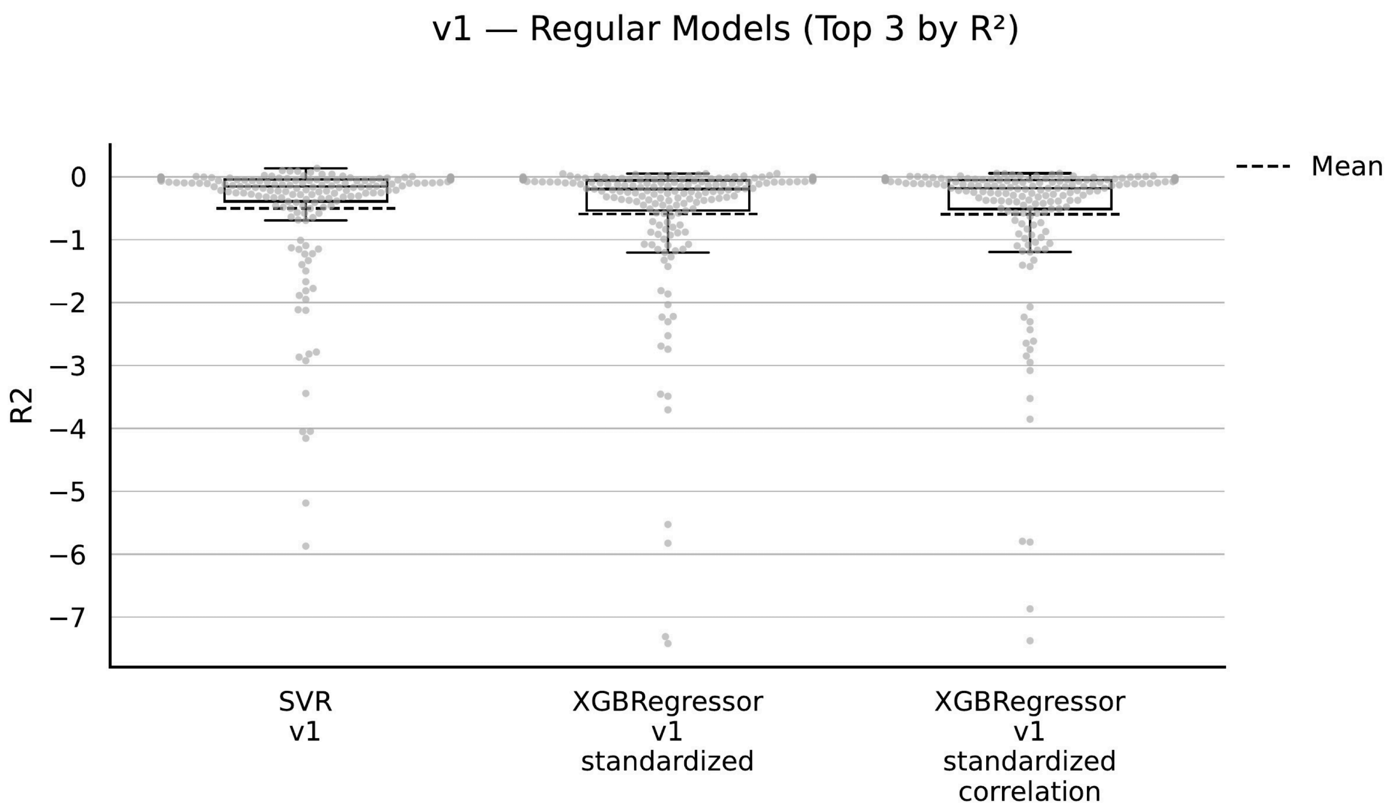

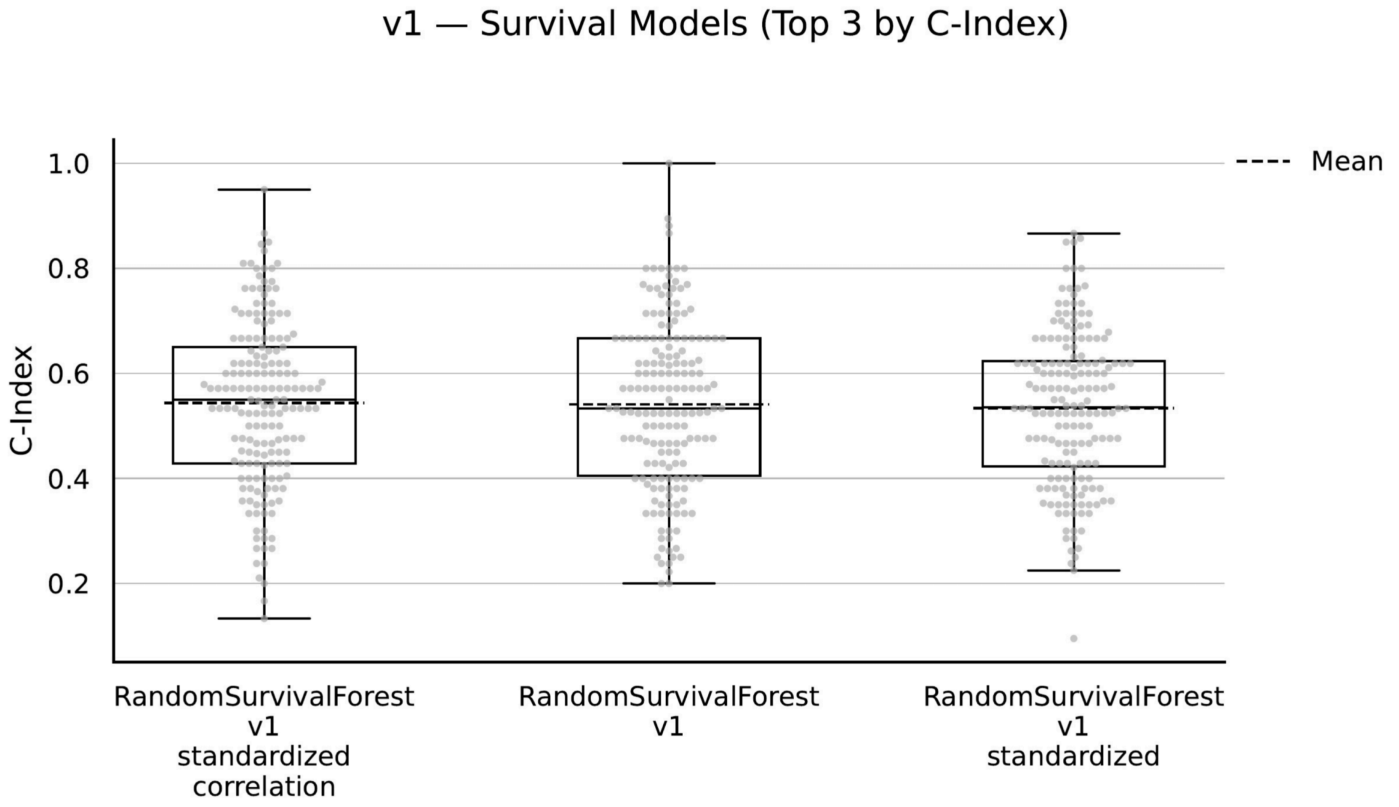

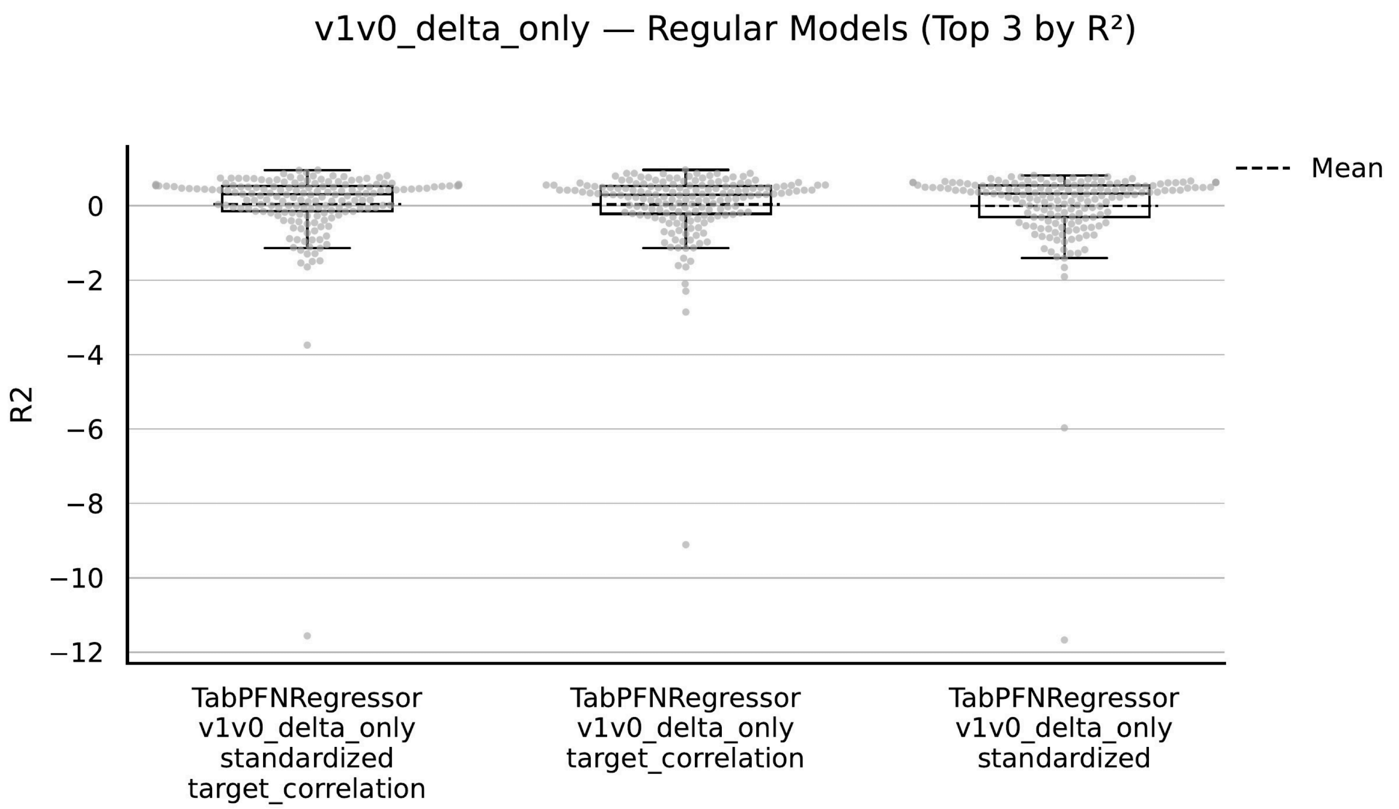

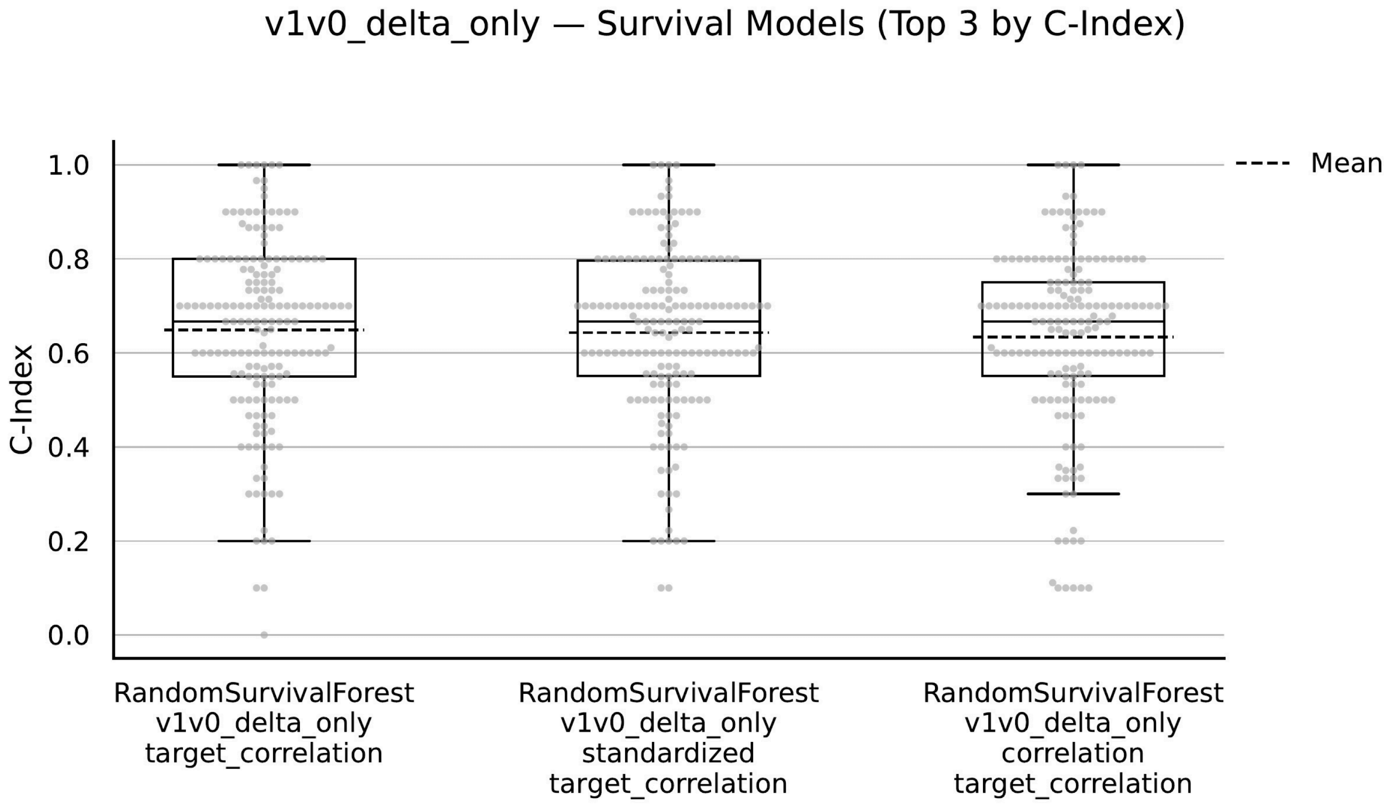
