## Supplementary material for "AURORA: Analysing and understanding responses to oncological regimens with artificial intelligence": Full Feature Ranking Lists

#### Legend - Explanation of Metrics.

Bootstrap-based metrics avg_bootstrap_freq: Fraction of bootstrap models where feature was selected.

avg_shap_boot: Mean importance across bootstrap runs using internal feature importance (tree split) or permutation importance Cross-validation metrics

avg_shap_cv: Mean SHAP importance across CV runs avg_cv_freq: Fraction of CV folds where feature was selected. aggregated_score: Weighted combination (0.3 * avg_cv + 0.3 * avg_boot + 0.2 * shap_cv(normalised) + 0.2 * shap_boot(normalised)).

Features with high selection frequency and SHAP are considered most stable and important.

| feature | avg_cv | avg_boot | avg_shap_cv | avg_shap_boot | aggregated_score |
| --- | --- | --- | --- | --- | --- |
| v0_Th17_CXCR3+ | 0.548 | 0.015 | 0.709 | 0.228 | 0.529 |
| v0_low_exhausted_CD4 | 0.768 | 0.026 | 0.889 | 0.07 | 0.499 |
| v0_CD8_cells | 0.621 | 0.006 | 0.348 | 0.064 | 0.322 |
| v0_EM_CD4 | 0.658 | 0.033 | 0.34 | 0.016 | 0.297 |
| v0_naive_CD8 | 0.677 | 0.005 | 0.309 | 0.021 | 0.292 |
| v0_Th17 | 0.605 | 0.017 | 0.084 | 0.077 | 0.273 |
| v0_CD56dim_CD16bright | 0.63 | 0.0 | 0.237 | 0.031 | 0.269 |
| v0_late_CD8 | 0.433 | 0.001 | 0.324 | 0.06 | 0.255 |
| v0_Th17_CXCR3- | 0.621 | 0.025 | 0.08 | 0.023 | 0.232 |
| v0_high_exhausted_CD4 | 0.562 | 0.017 | 0.166 | 0.008 | 0.218 |
| v0_T_cells_CD3 | 0.463 | 0.042 | 0.184 | 0.014 | 0.206 |
| v0_high_exhausted_CD8 | 0.456 | 0.0 | 0.299 | 0.0 | 0.204 |
| v0_EM_CD8 | 0.53 | 0.004 | 0.134 | 0.0 | 0.19 |
| v0_late_CD4 | 0.47 | 0.008 | 0.203 | 0.0 | 0.189 |
| v0_TE_CD4 | 0.552 | 0.017 | 0.039 | 0.0 | 0.179 |
| v0_TE_CD8 | 0.439 | 0.001 | 0.121 | 0.02 | 0.177 |
| v0_CD69_CD4 | 0.525 | 0.012 | 0.072 | 0.0 | 0.177 |
| v0_Lymph | 0.557 | 0.018 | 0.017 | 0.0 | 0.176 |
| v0_EMRA_CD4 | 0.52 | 0.017 | 0.066 | 0.0 | 0.176 |
| v0_CD4_cells | 0.428 | 0.033 | 0.054 | 0.029 | 0.176 |
| v0_naive_Treg_CD4 | 0.418 | 0.01 | 0.157 | 0.011 | 0.173 |
| v0_HLA_DR_CD4 | 0.489 | 0.01 | 0.097 | 0.0 | 0.172 |
| v0_CM_CD4 | 0.446 | 0.031 | 0.033 | 0.023 | 0.171 |
| v0_CM_CD8 | 0.49 | 0.003 | 0.072 | 0.0 | 0.164 |
| v0_CD56bright_CD16dim | 0.45 | 0.0 | 0.099 | 0.0 | 0.157 |
| v0_EMRA_CD8 | 0.461 | 0.003 | 0.053 | 0.0 | 0.151 |
| gender | 0.478 | 0.017 | 0.0 | 0.0 | 0.148 |
| v0_CD4_CD8_DP | 0.45 | 0.0 | 0.057 | 0.0 | 0.148 |
| v0_NK_like_T | 0.479 | 0.0 | 0.016 | 0.0 | 0.147 |
| v0_naive_CD4 | 0.461 | 0.016 | 0.01 | 0.0 | 0.146 |

### Feature Importance for patients_v0

| feature | avg_cv | avg_boot | avg_shap_cv | avg_shap_boot | aggregated_score |
| --- | --- | --- | --- | --- | --- |
| v0_memory_Treg_CD4 | 0.471 | 0.016 | 0.0 | 0.0 | 0.146 |
| v0_transitional_B | 0.462 | 0.01 | 0.017 | 0.0 | 0.145 |
| age | 0.46 | 0.007 | 0.022 | 0.0 | 0.145 |
| v0_B_cells | 0.452 | 0.003 | 0.031 | 0.0 | 0.144 |
| v0_interm_CD8 | 0.455 | 0.001 | 0.028 | 0.0 | 0.143 |
| v0_interm_CD4 | 0.467 | 0.01 | 0.0 | 0.0 | 0.143 |
| v0_Th2_CXCR3+ | 0.333 | 0.017 | 0.108 | 0.014 | 0.142 |
| v0_Th1 | 0.458 | 0.011 | 0.0 | 0.0 | 0.141 |
| v0_NK_cells | 0.455 | 0.0 | 0.015 | 0.0 | 0.14 |
| v0_Treg_CD8_CD127low | 0.464 | 0.0 | 0.0 | 0.0 | 0.139 |
| v0_early_CD8 | 0.442 | 0.002 | 0.012 | 0.0 | 0.136 |
| v0_naive_Treg_CD8 | 0.452 | 0.0 | 0.0 | 0.0 | 0.136 |
| v0_Th2_CXCR3- | 0.432 | 0.02 | 0.0 | 0.0 | 0.136 |
| v0_low_exhausted_CD8 | 0.4 | 0.001 | 0.072 | 0.0 | 0.136 |
| v0_class_switched_memory_B | 0.447 | 0.005 | 0.0 | 0.0 | 0.136 |
| v0_non_class_switched_memory_B | 0.444 | 0.006 | 0.0 | 0.0 | 0.135 |
| v0_TSCM_CD4 | 0.447 | 0.0 | 0.0 | 0.0 | 0.134 |
| v0_TSCM_CD8 | 0.447 | 0.0 | 0.0 | 0.0 | 0.134 |
| v0_CD69_CD8 | 0.387 | 0.0 | 0.032 | 0.0 | 0.123 |
| v0_HLA_DR_CD8 | 0.405 | 0.0 | 0.0 | 0.0 | 0.121 |
| v0_CD56_CD16 | 0.402 | 0.0 | 0.0 | 0.0 | 0.121 |
| v0_early_CD4 | 0.355 | 0.03 | 0.0 | 0.0 | 0.115 |
| v0_naive_B | 0.361 | 0.005 | 0.0 | 0.0 | 0.11 |
| v0_memory_Treg_CD8 | 0.322 | 0.0 | 0.019 | 0.0 | 0.101 |
| v0_Treg_CD4_CD127low | 0.282 | 0.011 | 0.039 | 0.0 | 0.097 |
| v0_Th2 | 0.287 | 0.015 | 0.0 | 0.0 | 0.091 |

### Feature Importance for patients_delta

| feature | avg_cv | avg_boot | avg_shap_cv | avg_shap_boot | aggregated_score |
| --- | --- | --- | --- | --- | --- |
| delta_v1_v0_naive_Treg_CD4 | 0.718 | 0.003 | 1.023 | 0.184 | 0.616 |
| delta_v1_v0_Th17_CXCR3+ | 0.777 | 0.014 | 0.181 | 0.084 | 0.364 |
| delta_v1_v0_non_class_switched_memory_B | 0.641 | 0.026 | 0.463 | 0.041 | 0.335 |
| delta_v1_v0_CD4_CD8_DP | 0.829 | 0.0 | 0.251 | 0.032 | 0.332 |
| delta_v1_v0_Th17 | 0.722 | 0.009 | 0.255 | 0.054 | 0.328 |
| delta_v1_v0_Th17_CXCR3- | 0.573 | 0.01 | 0.254 | 0.077 | 0.308 |
| delta_v1_v0_Th1 | 0.713 | 0.002 | 0.411 | 0.0 | 0.295 |
| delta_v1_v0_naive_CD8 | 0.709 | 0.003 | 0.241 | 0.026 | 0.289 |
| delta_v1_v0_EMRA_CD4 | 0.61 | 0.018 | 0.231 | 0.031 | 0.268 |
| delta_v1_v0_interm_CD4 | 0.798 | 0.002 | 0.043 | 0.0 | 0.248 |
| delta_v1_v0_CD56dim_CD16bright | 0.733 | 0.0 | 0.077 | 0.0 | 0.235 |
| delta_v1_v0_class_switched_memory_B | 0.754 | 0.005 | 0.029 | 0.0 | 0.233 |
| delta_v1_v0_Lymph | 0.572 | 0.017 | 0.168 | 0.021 | 0.233 |
| delta_v1_v0_low_exhausted_CD8 | 0.699 | 0.0 | 0.029 | 0.0 | 0.215 |
| delta_v1_v0_CD8_cells | 0.457 | 0.003 | 0.052 | 0.034 | 0.185 |
| delta_v1_v0_T_cells_CD3 | 0.49 | 0.034 | 0.062 | 0.014 | 0.184 |
| gender | 0.547 | 0.011 | 0.0 | 0.0 | 0.167 |
| delta_v1_v0_naive_Treg_CD8 | 0.483 | 0.0 | 0.088 | 0.0 | 0.162 |
| delta_v1_v0_Treg_CD4_CD127low | 0.52 | 0.004 | 0.0 | 0.0 | 0.157 |
| delta_v1_v0_TE_CD4 | 0.497 | 0.007 | 0.016 | 0.0 | 0.154 |
| delta_v1_v0_TSCM_CD4 | 0.447 | 0.0 | 0.097 | 0.0 | 0.153 |
| delta_v1_v0_HLA_DR_CD8 | 0.502 | 0.0 | 0.015 | 0.0 | 0.153 |
| delta_v1_v0_HLA_DR_CD4 | 0.484 | 0.004 | 0.023 | 0.0 | 0.151 |
| delta_v1_v0_TSCM_CD8 | 0.457 | 0.0 | 0.068 | 0.0 | 0.15 |
| delta_v1_v0_TE_CD8 | 0.436 | 0.0 | 0.094 | 0.0 | 0.149 |
| delta_v1_v0_Treg_CD8_CD127low | 0.453 | 0.0 | 0.06 | 0.0 | 0.148 |
| delta_v1_v0_CD69_CD4 | 0.465 | 0.005 | 0.038 | 0.0 | 0.148 |
| delta_v1_v0_high_exhausted_CD4 | 0.451 | 0.004 | 0.061 | 0.0 | 0.148 |
| delta_v1_v0_EM_CD8 | 0.486 | 0.001 | 0.003 | 0.0 | 0.147 |
| delta_v1_v0_CD56bright_CD16dim | 0.443 | 0.0 | 0.072 | 0.0 | 0.147 |

| feature | avg_cv | avg_boot | avg_shap_cv | avg_shap_boot | aggregated_score |
| --- | --- | --- | --- | --- | --- |
| delta_v1_v0_B_cells | 0.466 | 0.009 | 0.015 | 0.0 | 0.145 |
| delta_v1_v0_interm_CD8 | 0.457 | 0.0 | 0.018 | 0.0 | 0.141 |
| delta_v1_v0_CD69_CD8 | 0.402 | 0.0 | 0.035 | 0.01 | 0.138 |
| delta_v1_v0_high_exhausted_CD8 | 0.452 | 0.0 | 0.007 | 0.0 | 0.137 |
| delta_v1_v0_transitional_B | 0.45 | 0.004 | 0.005 | 0.0 | 0.137 |
| age | 0.455 | 0.004 | 0.0 | 0.0 | 0.137 |
| delta_v1_v0_naive_B | 0.378 | 0.006 | 0.104 | 0.0 | 0.136 |
| delta_v1_v0_late_CD4 | 0.436 | 0.004 | 0.003 | 0.0 | 0.133 |
| delta_v1_v0_late_CD8 | 0.416 | 0.0 | 0.031 | 0.0 | 0.131 |
| delta_v1_v0_EM_CD4 | 0.421 | 0.008 | 0.001 | 0.0 | 0.129 |
| delta_v1_v0_CM_CD8 | 0.39 | 0.001 | 0.06 | 0.0 | 0.129 |
| delta_v1_v0_low_exhausted_CD4 | 0.413 | 0.004 | 0.005 | 0.0 | 0.126 |
| delta_v1_v0_naive_CD4 | 0.369 | 0.01 | 0.046 | 0.0 | 0.123 |
| delta_v1_v0_EMRA_CD8 | 0.392 | 0.001 | 0.016 | 0.0 | 0.121 |
| delta_v1_v0_Th2_CXCR3+ | 0.373 | 0.007 | 0.002 | 0.0 | 0.114 |
| delta_v1_v0_early_CD4 | 0.32 | 0.015 | 0.06 | 0.0 | 0.112 |
| delta_v1_v0_early_CD8 | 0.299 | 0.001 | 0.047 | 0.01 | 0.11 |
| delta_v1_v0_CD56_CD16 | 0.36 | 0.0 | 0.0 | 0.0 | 0.108 |
| delta_v1_v0_Th2_CXCR3- | 0.337 | 0.01 | 0.0 | 0.0 | 0.104 |
| delta_v1_v0_memory_Treg_CD4 | 0.346 | 0.001 | 0.0 | 0.0 | 0.104 |
| delta_v1_v0_NK_cells | 0.333 | 0.0 | 0.014 | 0.0 | 0.103 |
| delta_v1_v0_Th2 | 0.332 | 0.012 | 0.0 | 0.0 | 0.103 |
| delta_v1_v0_CD4_cells | 0.307 | 0.014 | 0.032 | 0.0 | 0.102 |
| delta_v1_v0_memory_Treg_CD8 | 0.317 | 0.0 | 0.021 | 0.0 | 0.099 |
| delta_v1_v0_NK_like_T | 0.326 | 0.0 | 0.004 | 0.0 | 0.098 |
| delta_v1_v0_CM_CD4 | 0.3 | 0.013 | 0.0 | 0.0 | 0.094 |
