## Supplementary material for "AURORA: Analysing and understanding responses to oncological regimens with artificial intelligence": Model Hyperparameter Grids

**Table D.** XGBoost (XGB)	**Table E.** Random Forest (RF)

| **Parameter** | **Values** |
| --- | --- |
| *n*estimators | {3, 5, 10, 20, 50} |
| max depth | {2, 3} |
| min samples split | {2, 4} |
| min samples leaf | {1, 2} |
| max features | {"sqrt", "log2"} |
| bootstrap | True |

| **Parameter** | **Values** |
| --- | --- |
| *n*estimators | {3, 5, 10, 20, 35, 50} |
| learning rate | {0.05, 0.1} |
| max depth | {2, 3} |
| subsample | {0.8, 1.0} |
| colsample bytree | {0.8, 1.0} |
| reg alpha | {0.5, 1} |
| reg lambda | {0.5, 1, 2} |

**Table F.** Ridge Regression (Ridge)	**Table G.** Lasso Regression (Lasso)

| **Parameter** | **Values** |
| --- | --- |
| *Α* | {0.01, 0.1, 1.0, 10.0, 50, 100.0} |
| Solver | "auto" |
| fit intercept | {True, False} |
| max iter | 100 |

| **Parameter** | **Values** |
| --- | --- |
| *α* | {0.1, 0.001, 0.01, 0.1, 1.0} |
| max iter | 100 |
| fit intercept | {True, False} |

**Table H.** Support Vector Regression (SVR)	**Table I.** Tabular Prior-Data Fitted Network (TabPFN)

| **Parameter** | **Values** |
| --- | --- |
| *C* | {0.1, 1, 10, 100} |
| *ϵ* | {0.01, 0.1, 1} |
| kernel | {"linear", "rbf"} |
| *γ* | {"scale", 0.1, 0.01} |

| **Parameter** | **Values** |
| --- | --- |
| n estimators | {1, 3, 5, 10} |

**Table J.** Nearest Neighbors Regressor (NNR)	**Table K.** Random Survival Forest (RSF)

| **Parameter** | **Values** |
| --- | --- |
| *n*estimators | {5, 10, 20, 30, 50} |
| max depth | {1, 2, 3} |
| min samples split | {1, 2, 3, 4} |
| min samples leaf | {1, 2} |
| max features | {"sqrt", "log2"} |
| bootstrap | True |

| **Parameter** | **Values** |
| --- | --- |
| *n*neighbors | {2, 3, 4} |
| weights | {"uniform", "distance"} |
| algorithm | "auto" |
| *p* | {1, 2} |

**Table L.** CoxNet (Elastic-Net Cox)	**Table M.** Lifelines CoxPH (LLC)

| **Parameter** | **Values** |
| --- | --- |
| alphas | {0.01, 0.1, 1.0, 10, 50} |
| l1 ratio | {0.1, 0.25, 0.5, 0.75, 1.0} |
| fit baseline model | True |

| **Parameter** | **Values** |
| --- | --- |
| penalizer | 1 |

**Table N.** Survival SVM (SVRSurv)

| **Parameter** | **Values** |
| --- | --- |
| *α* | {0.001, 0.01, 0.1, 0.5, 1.0} |
| tol | {1e-4, 1e-3} |
| rank ratio | {0.0, 0.5, 0.95} |
| fit intercept | {False, True} |
| optimizer | {"avltree", "rbtree", "direct-count"} |
| max iter | {1, 2500, 5, 10, 100} |
